# Unmeasured but Not Unbiased: The Missingness Demographic Leakage Audit (MDLA) for Calibration-Aware Fairness Evaluation in Critical Care Mortality Prediction

**DOI:** 10.64898/2026.05.01.26352193

**Authors:** Krutarth Patel, Phanindra Beedala

**Affiliations:** Department of Interoperability, Humana Inc., Louisville, KY, USA; Independent Researcher, Louisville, KY, USA

**Keywords:** Intensive Care Units, Clinical Prediction Models, Algorithmic Fairness, Model Calibration, Hospital Mortality, Missing Not at Random

## Abstract

**Objective:** Clinical prediction models trained on electronic health records are routinely evaluated for fairness on observed feature values, but the informativeness of which measurements are absent remains unaudited. We developed the Missingness Demographic Leakage Audit (MDLA), a reproducible four-step informatics framework that tests whether patterns of clinical measurement absence function as latent demographic proxies — constituting a bias pathway invisible to standard fairness audits.

**Materials and Methods:** We applied MDLA across development (MIMIC-IV v2.2; n=50,827; mortality 10.2%) and external validation (eICU-CRD v2.0; n=137,773; mortality 9.5%) cohorts following TRIPOD+AI standards. XGBoost, random forest, and logistic regression were trained on 43 clinical features and 44 binary missingness indicators. MDLA quantified demographic predictability from missingness alone, tested feature-level associations with Bonferroni correction, and verified model reliance via ablation. A calibration-aware fairness audit evaluated five criteria across four demographic axes; six post-hoc recalibration strategies were compared on a fairness-utility Pareto frontier.

**Results:** Missingness indicators alone predicted racial group membership above chance (AUROC=0.543; 95% CI, 0.540–0.546), with 18 of 43 features showing Bonferroni-significant race-missingness associations (all Cramér’s V<0.10). Ablation confirmed model reliance: adding missingness indicators increased racial AUROC disparity by 10.7% (0.063 to 0.069) without improving global performance. XGBoost achieved AUROC=0.910 internally (AUROC=0.799 on external validation). Global Platt recalibration reduced overall calibration error by 94% and maximum racial calibration error by 51%, with zero AUROC loss and successful parameter transfer to external validation without retraining.

**Conclusion:** MDLA provides a structured, reproducible protocol for detecting missingness-encoded demographic signals prior to model deployment. Applied across 188,600 ICU patient-stays from two institutionally diverse databases, it identified a statistically confirmed but subtle bias pathway undetectable by standard fairness audits. Missingness-aware auditing and calibration-aware evaluation should be integrated into clinical AI validation pipelines.

## 1. INTRODUCTION

Intensive care units (ICUs) account for approximately 20% of Medicare hospitalizations in the United States, with in-hospital mortality rates of 8%–19% and an estimated 500,000 deaths annually [1,2]. Accurate mortality prediction is central to ICU decision-making — informing triage, goals-of-care discussions, and resource allocation where timely decisions carry life-or-death consequences [3,4]. Machine learning models trained on electronic health records have demonstrated strong discriminative performance for this task, with AUROC values routinely exceeding 0.85 in single-center studies [5–8]. However, population-level performance masks a critical concern: models performing well on average may perform inequitably across demographic subgroups, compromising reliability and generalizability across the deployment populations these models are intended to serve.

Obermeyer and colleagues [9] compellingly demonstrated that a widely deployed risk algorithm produced racial bias through a biased proxy outcome. Subsequent work identified analogous disparities in critical care and clinical imaging AI [10–12], motivating increasing calls for formal fairness auditing before deployment [13,14]. Existing audits have focused predominantly on discriminative performance disparities, despite calibration fairness being equally critical for clinical deployment: a model well-calibrated globally but poorly calibrated within minority subgroups systematically misestimates absolute mortality risk, potentially contributing to under-triage or inappropriate withdrawal of care [15,16].

Despite this progress, existing frameworks share a fundamental blind spot: they examine disparities in what models predict from observed values but ignore disparities in which measurements are absent. Missing data is pervasive in critical care — laboratory values, physiologic measurements, and neurological assessments are frequently absent for reasons that may reflect differences in ordering practices, resource availability, and structural inequities tied to race, insurance status, and socioeconomic position [17,18]. Standard approaches treat missingness as a nuisance requiring imputation [19,20]. An undercharacterized possibility is that absence patterns carry statistically significant demographic information, and that models may rely on this signal without explicit awareness — constituting an upstream bias pathway invisible to standard fairness audits. This pathway has not been ablation-verified across independent ICU datasets, and no reproducible informatics framework exists to audit it as a standard component of clinical AI validation.

To address this gap, we developed the Missingness Demographic Leakage Audit (MDLA) — a structured, reproducible four-step protocol that explicitly tests whether absence patterns function as latent demographic proxies, designed for integration into calibration-aware clinical AI validation pipelines. We apply MDLA within a calibration-aware fairness audit following TRIPOD+AI standards [21,22], across two databases representing 188,600 patient-stays. All analytical decisions — including preprocessing, threshold selection, and recalibration strategies — were derived exclusively from the development cohort and applied without modification to external validation, consistent with prospective deployment conditions.

The objectives are: (1) to characterize demographic information in missingness patterns using MDLA and assess model reliance via ablation; (2) to conduct a multi-criteria fairness audit across five criteria covering race and ethnicity, sex, insurance status, age, and ICU type; (3) to compare post-hoc recalibration strategies using a fairness-utility Pareto frontier; and (4) to quantify the generalization gap between development and external validation cohorts using distributional shift analysis.

### Statement of Significance

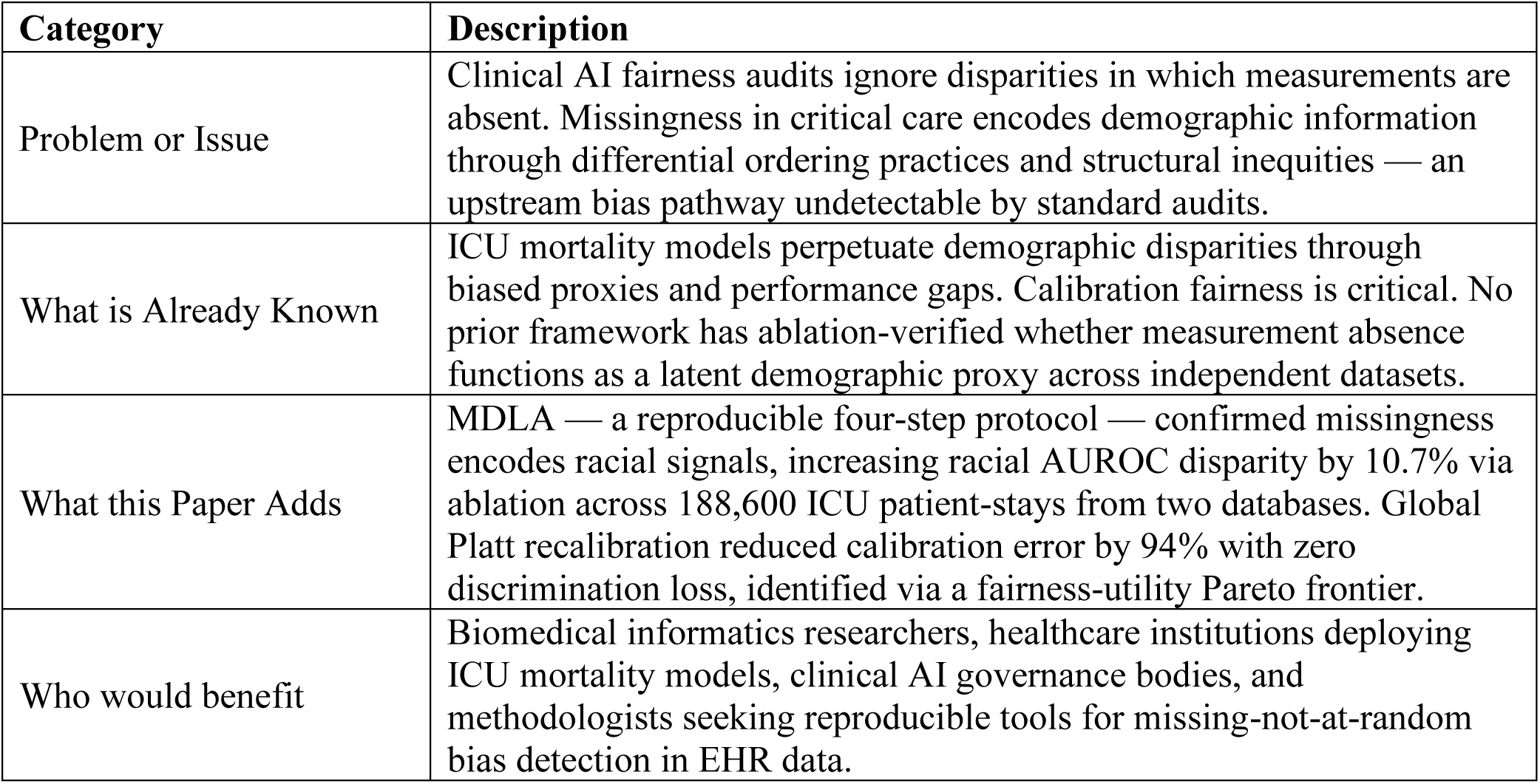

| Category | Description |
| --- | --- |
| Problem or Issue | Clinical AI fairness audits ignore disparities in which measurements are absent. Missingness in critical care encodes demographic information through differential ordering practices and structural inequities — an upstream bias pathway undetectable by standard audits. |
| What is Already Known | ICU mortality models perpetuate demographic disparities through biased proxies and performance gaps. Calibration fairness is critical. No prior framework has ablation-verified whether measurement absence functions as a latent demographic proxy across independent datasets. |
| What this Paper Adds | MDLA — a reproducible four-step protocol — confirmed missingness encodes racial signals, increasing racial AUROC disparity by 10.7% via ablation across 188,600 ICU patient-stays from two databases. Global Platt recalibration reduced calibration error by 94% with zero discrimination loss, identified via a fairness-utility Pareto frontier. |
| Who would benefit | Biomedical informatics researchers, healthcare institutions deploying ICU mortality models, clinical AI governance bodies, and methodologists seeking reproducible tools for missing-not-at-random bias detection in EHR data. |

## 2. METHODS

### 2.1 Study Design and Reporting

We conducted a retrospective cohort study using two independently collected critical care databases. All preprocessing parameters, imputation statistics, and operating thresholds were derived exclusively from the development training set and applied without modification to subsequent splits, consistent with prospective-style external validation. This study follows the TRIPOD+AI reporting standards (**Supplementary Table S8**) [21,22]. Analyses were conducted in Python 3.11 using scikit-learn 1.4.2 [23], XGBoost 2.0 [24], statsmodels 0.14, and SHAP 0.44 [25,26]. Code and outputs are publicly available at [mdla-icu-fairness-audit].

### 2.2 Data Sources and Cohort Construction

#### Development cohort

We extracted data from MIMIC-IV v2.2 [27], a single-center critical care database from Beth Israel Deaconess Medical Center (Boston, MA). Eligible patients met all of the following criteria: (1) age ≥18 years at ICU admission; (2) ICU length of stay ≥4 hours; (3) non-missing in-hospital mortality outcome; and (4) first ICU stay per hospital admission and first hospital admission per patient. These criteria yielded 50,827 ICU stays (in-hospital mortality 10.2%).

#### External validation cohort

The eICU Collaborative Research Database v2.0 [28], aggregating 208 ICUs across the United States, provided external validation. Identical inclusion criteria yielded 137,773 ICU stays (mortality 9.5%). Both databases are accessed through PhysioNet [29] under credentialed data use agreements.

#### Demographic harmonization

Race and ethnicity were harmonized to five groups (White, Black, Hispanic, Asian, Other). Insurance was mapped to Medicare, Medicaid, and Other/Unknown — a known limitation of MIMIC-IV v2.2, which removed granular private insurance categorization from MIMIC-III. Insurance fairness analyses were restricted to the development cohort, as eICU does not collect insurance data. ICU type was harmonized to six categories (MICU, SICU, CCU, NICU, TSICU, Other). Age was grouped into four strata: 18–44, 45–64, 65–74, and ≥75 years.

### 2.3 Feature Extraction

All features were extracted from a fixed 24-hour window beginning at ICU admission, summarized as minimum, maximum, and mean per stay. Twenty vital sign features were derived from heart rate, systolic blood pressure, mean arterial pressure, respiratory rate, SpO₂, temperature, and GCS total. Twenty-two laboratory values were initially defined; troponin was excluded due to 100% training missingness, yielding 21 laboratory predictors. Two clinical composites were included: 24-hour urine output and Elixhauser comorbidity count [30,31] (MIMIC-IV only; 100% missing in eICU). The final predictor set comprised 43 features (20 vital + 21 laboratory + 2 clinical). Laboratory values were filtered using clinically defined plausibility bounds (e.g., creatinine 0.1–30 mg/dL, pH 6.5–8.0).

### 2.4 Missingness Demographic Leakage Audit (MDLA)

Prior to imputation, 44 binary missingness indicators were constructed — one per initially defined feature including excluded troponin — where miss_X = 1 if feature X was absent. While imputation proceeded under a missing-at-random (MAR) assumption [19,32], MDLA explicitly tested whether this held with respect to demographic variables; significant demographic predictability from missingness alone would constitute evidence toward missing-not-at-random (MNAR) structures. MDLA comprised four steps:

#### Step 1 — Indicator construction

Binary missingness indicators were constructed before any imputation.

#### Step 2 — Demographic predictability testing

A multinomial logistic regression classifier was trained on missingness indicators alone (no clinical values) to predict racial group membership and insurance type, using 5-fold cross-validated macro-averaged one-versus-rest AUROC. Confidence intervals were computed analytically using the Hanley-McNeil variance formula [33].

#### Step 3 — Feature-level association quantification

Chi-square tests of independence were computed between each missingness indicator and racial group; the troponin indicator was excluded from this step because miss_troponin = 1 for all training observations (a constant provides no discriminative information for chi-square testing), yielding 43 testable indicators. Effect size was quantified using Cramér’s V. All p-values were Bonferroni-corrected for the 43 tests conducted, with significance defined at corrected p < 0.05.

#### Step 4 — Model reliance ablation

XGBoost was retrained on the augmented feature matrix comprising the original 43 predictors concatenated with all 44 missingness indicators (87 features total). All 44 indicators — including miss_troponin — were retained in the ablation matrix; the exclusion of miss_troponin in Step 3 applied only to chi-square association testing, not to model training, where it contributes a valid structural signal. The change in racial AUROC disparity between the standard and augmented models quantified the extent to which the model relies on demographic signals in missingness patterns.

### 2.5 Data Partitioning and Preprocessing

The development cohort was partitioned into training (70%, n=35,578), internal validation (15%, n=7,624), and internal test (15%, n=7,625) sets using stratified random splits preserving mortality prevalence. Median imputation parameters were fit on the training set only and applied to all subsequent partitions; this strategy was retained to isolate the effect of missingness indicators as explicit features in MDLA Step 4, with the understanding that confirmed MNAR signals are themselves an argument for more sophisticated imputation in future deployments (Section 4.5). Min-max scaling [0, 1] was applied to logistic regression inputs; tree-based models received unscaled features. All random operations used a fixed seed (42).

### 2.6 Model Development

Three classifiers were developed: logistic regression (LR), random forest (RF) [34], and XGBoost (XGB) [24]. Hyperparameters were selected via 5-fold stratified grid search optimizing AUROC; full grid specifications are provided in **Supplementary Table S1**. Operating thresholds were selected at the Youden index optimum on the internal validation set for comparative evaluation; these thresholds are not intended as clinical deployment recommendations.

### 2.7 Performance Evaluation

Global performance was assessed using AUROC (1000-iteration bootstrap 95% CIs), AUPRC, Brier score [35], DeLong’s test for pairwise AUROC comparisons [36], expected calibration error (ECE, 10 uniform bins) [37], and calibration slope and intercept via logistic recalibration on logit-transformed predictions [16]. Classification metrics were computed at the Youden threshold.

### 2.8 Formal Fairness Assessment

Five fairness criteria were operationalized [38–42]: equalized odds (maximum TPR and FPR differences), equal opportunity (maximum TPR difference), demographic parity (maximum positive prediction rate difference), predictive parity (maximum PPV difference), and calibration fairness (maximum group ECE). Criteria were evaluated across race and ethnicity, sex, insurance type, and ICU type. Age-group stratification was not evaluated as a primary fairness axis because age is a clinically legitimate predictor of mortality; observed performance differences across age strata reflect prognostic differentiation rather than inequitable treatment of a protected class. Bootstrap CIs (1000 iterations) were computed for pairwise AUROC, TPR, and ECE gaps. Groups with n<30 in the test partition were excluded. Fairness-performance incompatibility was evaluated empirically following Chouldechova’s impossibility theorem [39].

### 2.9 Post-Hoc Recalibration

Six recalibration strategies were evaluated on XGBoost. All calibrators were fit on the internal validation set and evaluated on the held-out internal test set and external validation set separately: (1) uncalibrated reference; (2) global Platt scaling [43] via logistic regression on logit-transformed validation predictions; (3) group-conditional Platt scaling by race, with groups below n=50 in the validation set falling back to global calibration; (4) group-conditional Platt scaling by insurance type; (5) isotonic regression [44]; and (6) fairness-aware temperature scaling [45], which rescales pre-calibration logits by a learned scalar T (calibrated probability = σ(logit / T)) optimized to minimize ECE + λ·MaxGroupECE (λ=0.5) via bounded scalar search on the validation set; λ=0.5 was selected to weight overall and group-level calibration error equally as a pre-specified default; unlike Platt scaling, temperature scaling adjusts logit spread only and cannot correct intercept-level shifts [45]. Strategies were compared using a fairness-utility Pareto frontier [46] plotting overall ECE against racial calibration gap. Decision curve analysis was performed for each model at the Youden-index threshold [47].

### 2.10 Domain Shift Analysis

To characterize the generalization gap between internal and external validation, Kolmogorov-Smirnov two-sample tests were computed between development and external validation cohort distributions for 41 of the 43 predictors; Elixhauser comorbidity count and 24-hour urine output were excluded from distributional comparison because both features are 100% missing in the eICU cohort, rendering a two-sample test undefined. Results are reported descriptively to identify primary drivers of distributional shift; p-values are reported without multiple testing correction given the exploratory nature of this analysis.

### 2.11 Ethical Considerations

Both databases are de-identified and publicly available through PhysioNet [29] under credentialed data use agreements. This study used secondary data only, did not involve patient contact, and did not use or retain identifiable information. Institutional review board approval was not required.

## 3. RESULTS

### 3.1 Cohort Characteristics

The development cohort comprised 50,827 ICU stays (in-hospital mortality 10.2%; mean age 63.5 years, SD 17.2; 44.2% female). Racial and ethnic composition was predominantly White (67.2%), followed by Other/Unknown (17.7%), Black (9.1%), Hispanic (3.4%), and Asian (2.6%). Insurance was categorized as Medicare (42.8%), Other/Unknown (50.0%), and Medicaid (7.1%), with no granular private insurance designation available in MIMIC-IV v2.2. The external validation cohort comprised 137,773 ICU stays from 208 institutions (mortality 9.5%; mean age 63.3 years, SD 17.2; 45.9% female), with a higher proportion of White patients (77.4%) and broader geographic distribution. Full cohort characteristics are presented in **Table 1**.

**Table 1:** Cohort Characteristics. BIDMC = Beth Israel Deaconess Medical Center. MIMIC-IV v2.2 does not provide granular private insurance categorization; ‘Other/Unknown’ includes commercially insured and uninsured patients. All partition splits stratified by outcome prevalence. eICU-CRD held entirely out until final evaluation; no preprocessing parameters derived from it. Vital features: heart rate, systolic BP, MAP, respiratory rate, SpO2, temperature, GCS total (min/max/mean each = 20 features). Laboratory features: creatinine, BUN, lactate, hemoglobin, hematocrit, platelets, WBC, sodium, potassium, bicarbonate, chloride, glucose, PaO2, PaCO2, pH, ALT, AST, bilirubin, albumin, INR, PTT (troponin excluded due to 100% training missingness = 21 features). Clinical composites: 24-hour urine output, Elixhauser comorbidity count.

| Variable | Development Cohort<br>(MIMIC-IV v2.2) | External Validation (eICU-CRD v2.0) |
| --- | --- | --- |
| **Study Design** |  |  |
| Role | Development (train/val/test) | External validation only |
| ICU sites | 1 (BIDMC, Boston MA) | 208 (multi-site, USA) |
| Database version | MIMIC-IV v2.2 | eICU-CRD v2.0 |
| **Partition Sizes** |  |  |
| Training set, n | 35,578 | — |
| Internal validation, n | 7,624 | — |
| Internal test, n | 7,625 | — |
| External validation, n | — | 137,773 |
| **Demographics** |  |  |
| ICU stays, n | **50,827** | **137,773** |
| Age, mean (SD), years | 63.5 (17.2) | 63.3 (17.2) |
| Female, % | 44.2 | 45.9 |
| **Race and Ethnicity, %** |  |  |
| White | 67.2 | 77.4 |
| Black | 9.1 | 10.5 |
| Hispanic | 3.4 | 3.8 |
| Asian | 2.6 | 1.7 |
| Other / Unknown | 17.7 | 6.7 |
| **Insurance, %** |  |  |
| Medicare | 42.8 | — |
| Medicaid | 7.1 | — |
| Other / Unknown | 50 | — |
| Not collected | — | Yes |
| **Outcomes** |  |  |
| In-hospital mortality, % | **10.2** | **9.5** |
| Class imbalance ratio<br>(neg:pos) | 8.8 : 1 | 9.5 : 1 |
| **Feature Set** |  |  |
| Total predictors (after<br>exclusions) | 43 | 43 |
| Vital sign features | 20 | 20 |
| Laboratory features | 21 (troponin excluded) | 21 |
| Clinical composite features | 2 | 2 |
| Missingness indicators | 44 | 44 |
| Elixhauser comorbidity | Available (ICD-10) | 100% missing (no ICD) |

### 3.2 MDLA Steps 1–3: Missingness Patterns Carry Statistically Significant Demographic Information

Differential missingness rates across demographic subgroups are visualized in **Figure 1**.

**Figure 1.**
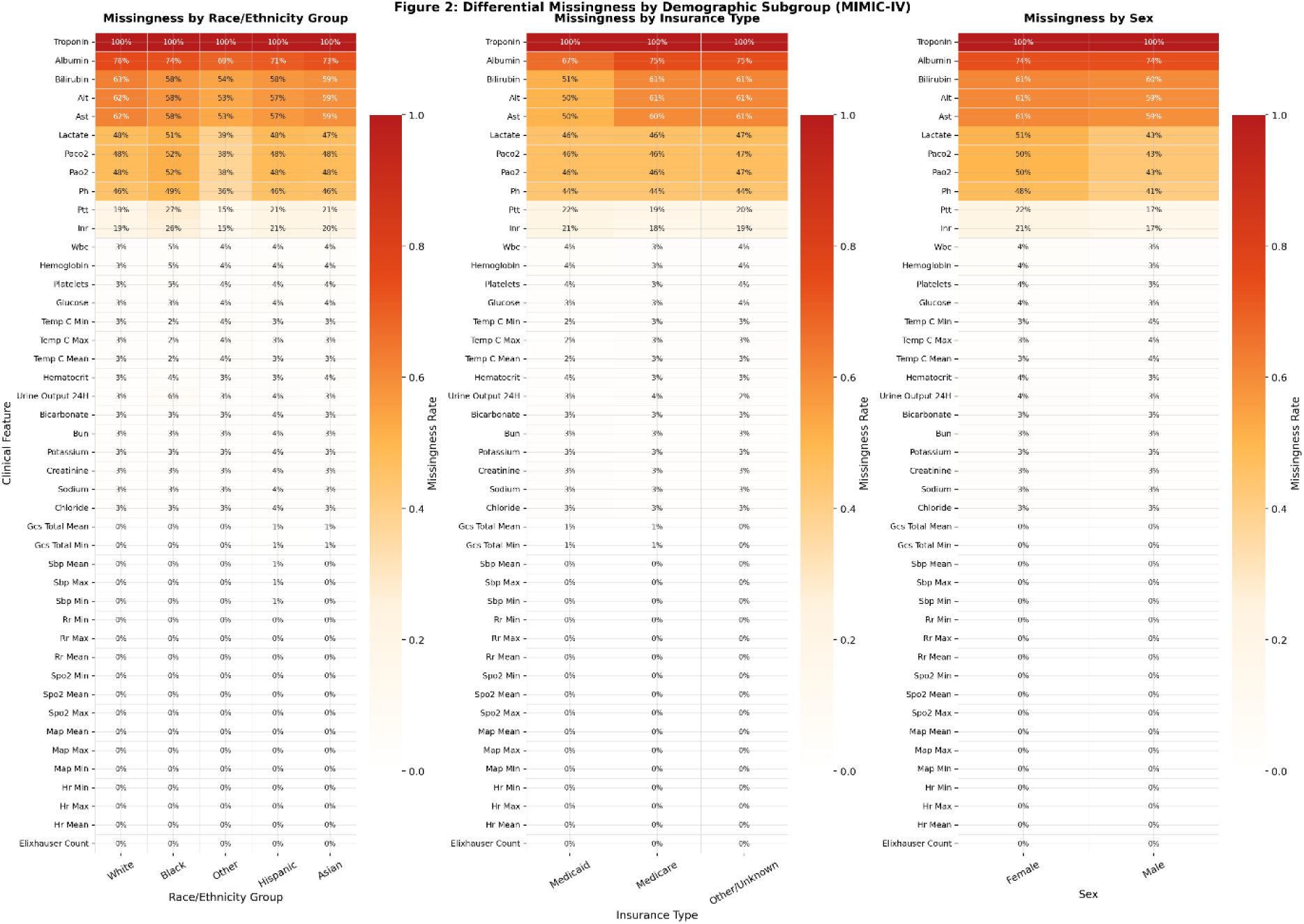
Differential Missingness Rates Across Demographic Subgroups in the Development Cohort. Heatmap of missingness rates for all 43 predictors stratified by race/ethnicity (left), insurance type (center), and sex (right). Features ordered by overall missingness rate. Arterial blood gas measurements show the largest racial gaps (up to 14 percentage points). Development cohort; n=50,827.

Three findings emerged from MDLA Steps 1–3: arterial blood gas measurements showed the largest racial gaps, missingness patterns carried statistically confirmed demographic predictability above chance, and all effect sizes remained small, indicating a subtle rather than severe encoding signal.

Arterial blood gas measurements (PaCO₂, PaO₂, pH) showed the largest racial missingness gaps (overall missingness: 46.4%, 46.4%, 44.2%). Black patients showed 51.6% vs 47.9% White missingness for PaCO₂ and PaO₂ (gap=0.140 for both; both analytes co-measured from a single arterial draw). Coagulation markers showed structured gaps: PTT gap=0.118 (Black 27.2% vs White 19.1%); INR gap=0.114. Sex-stratified missingness was modest across all features (**Supplementary Table S2**).

MDLA Step 2 demonstrated that missingness patterns alone predicted demographic group membership above chance (**Figure 2**).

**Figure 2.**
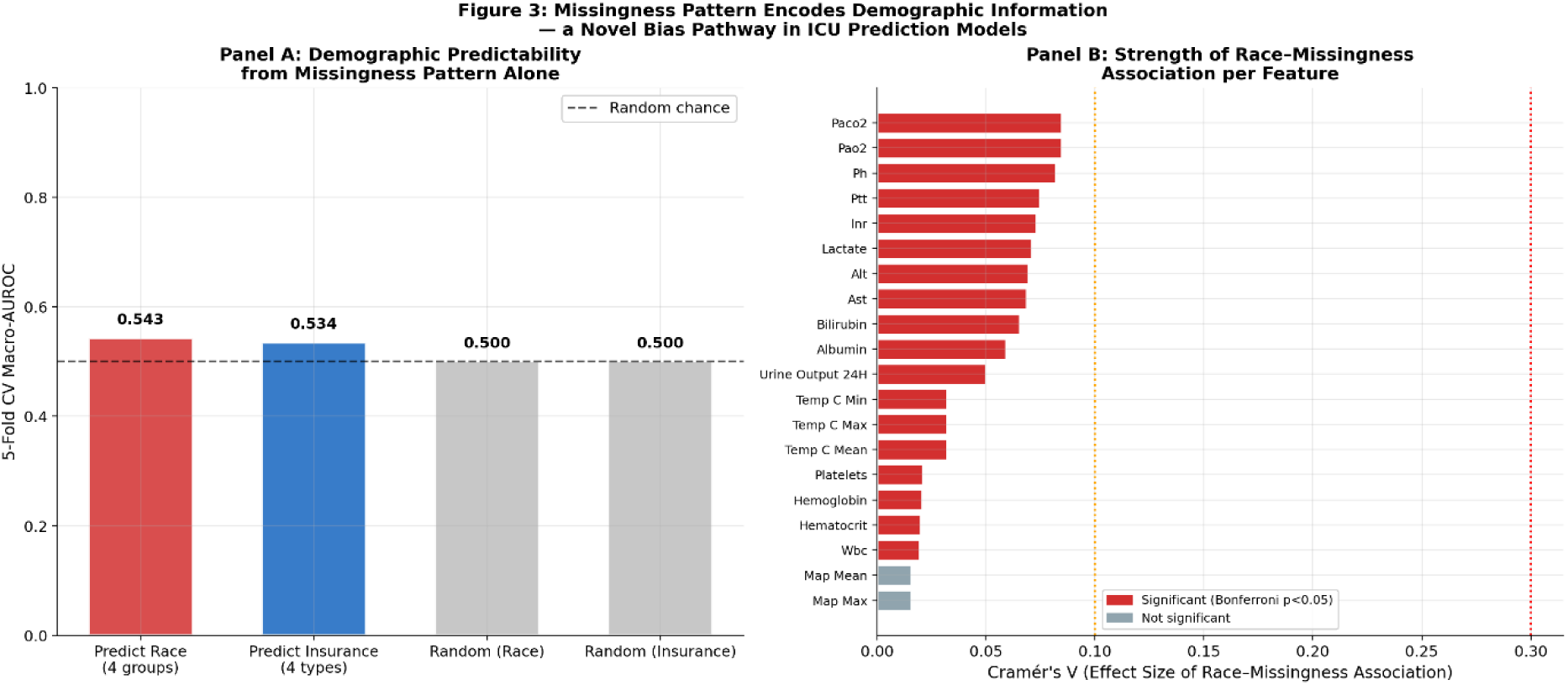
MDLA Steps 2–3: Demographic Predictability from Missingness Patterns Alone. Panel A: Cross-validated AUROC for predicting racial group and insurance type from 44 binary missingness indicators with no clinical values; dashed line = chance (0.500). Panel B: Cramér’s V effect sizes per feature; red = Bonferroni-significant (18/43 features); all V<0.10 indicating a small but confirmed signal. Development cohort; Bonferroni correction across 43 tests.

A logistic regression trained exclusively on 44 binary missingness indicators predicted racial group membership (AUROC=0.543; 95% CI: 0.540–0.546) and insurance type (AUROC=0.534; 95% CI: 0.531–0.539), both with CIs excluding 0.500 (**Supplementary Table S4**). MDLA Step 3 confirmed feature-level associations: 18 of 43 features showed Bonferroni-significant race-missingness associations (corrected p<0.05; **Supplementary Table S3**), with the largest effects for PaCO₂ (Cramér’s V=0.085), PaO₂ (V=0.085), and pH (V=0.082). All Cramér’s V values were below 0.10 (**Figure 2, Panel B**) — statistically significant but small, consistent with a subtle rather than overt encoding signal.

### 3.3 MDLA Step 4: Evidence of Model Reliance on Missingness-Encoded Demographic Signals

Ablation indicated that XGBoost models may rely on these patterns. Retraining on the augmented matrix (43 predictors + 44 missingness indicators) increased racial AUROC disparity from 0.063 to 0.069 (+10.7%) while global AUROC remained stable (0.910 vs 0.912), indicating differential model weighting of demographic signals rather than overall performance gain (**Table 2**).

**Table 2:**
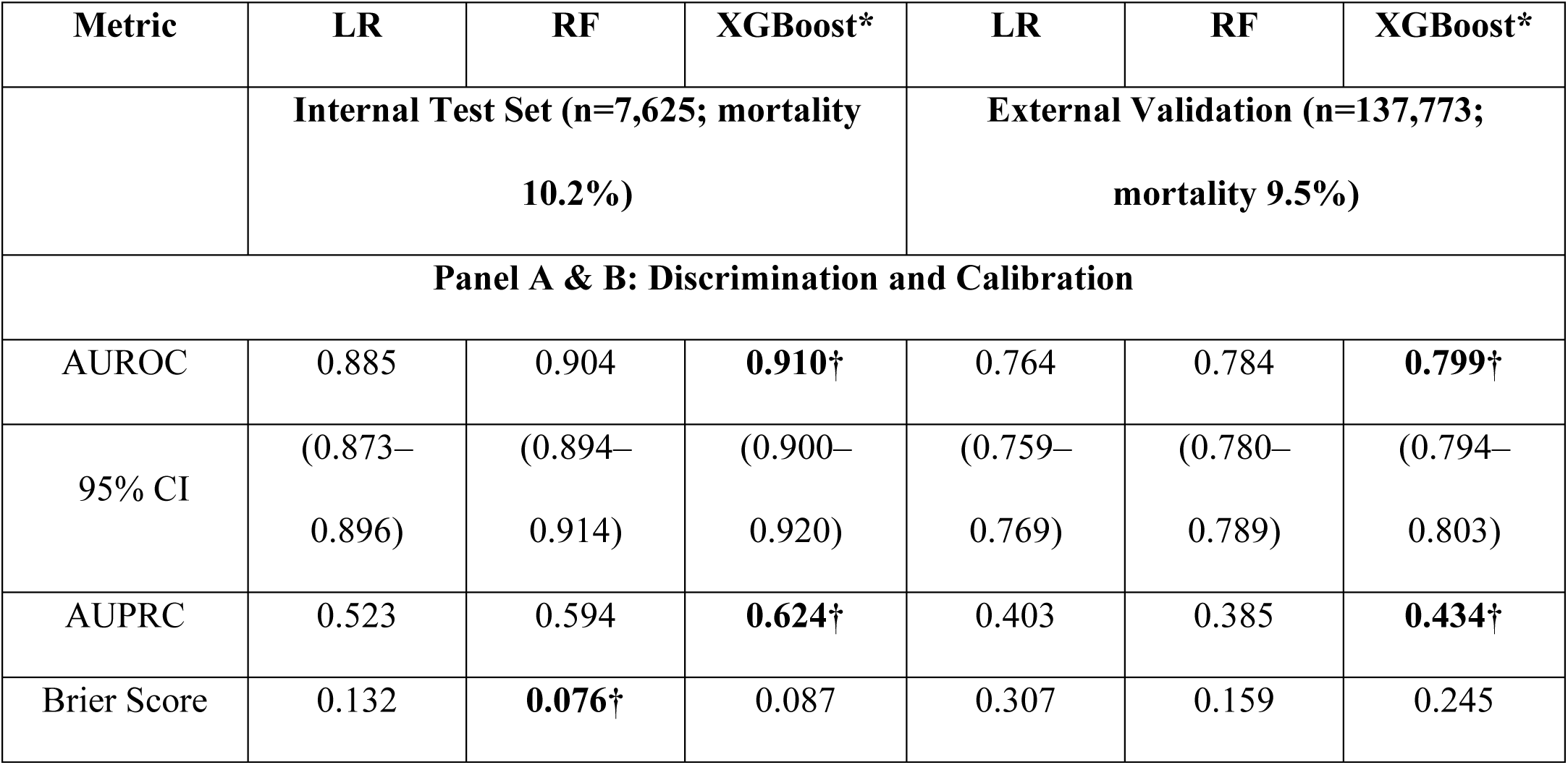
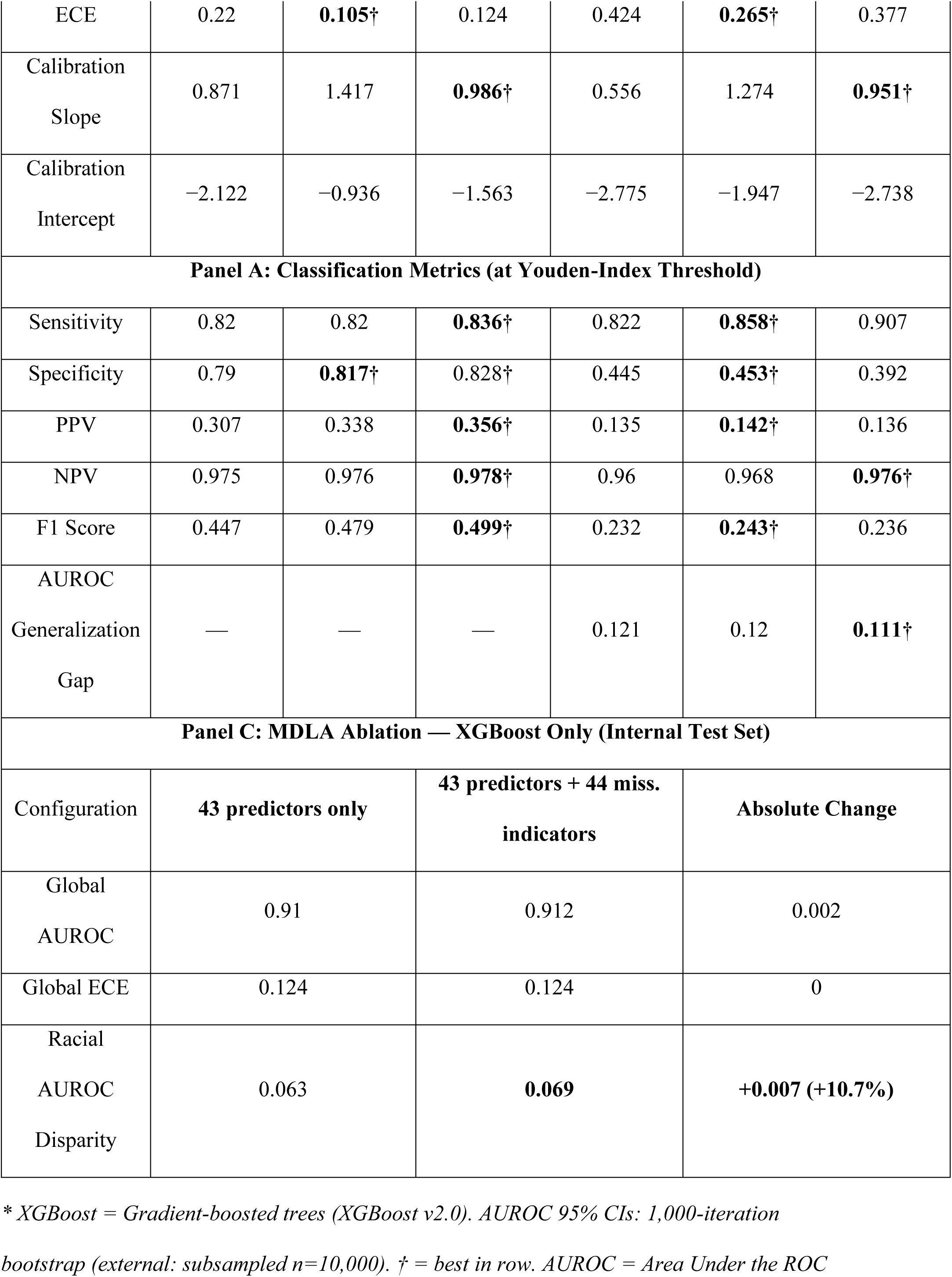
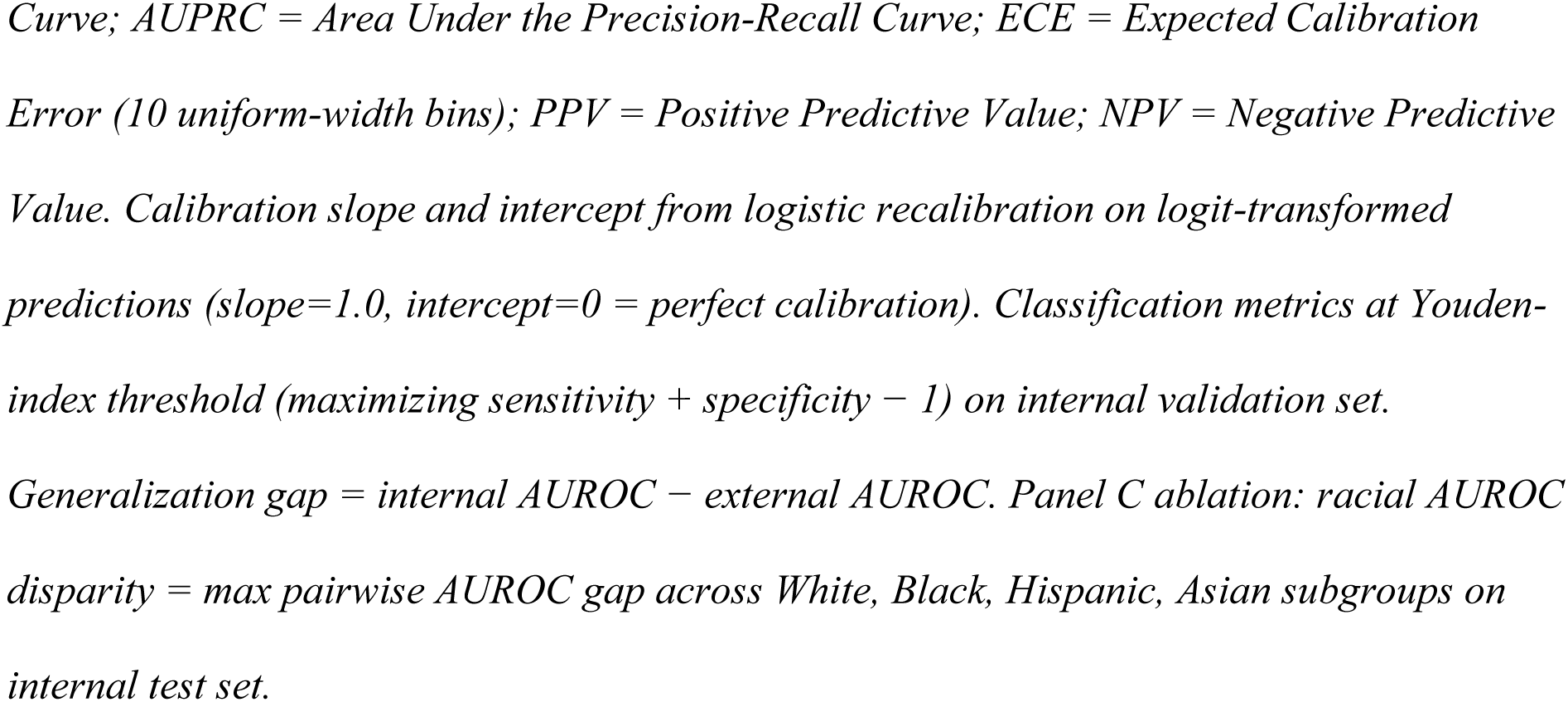
Model Performance — Internal Test Set and External Validation. * XGBoost = Gradient-boosted trees (XGBoost v2.0). AUROC 95% CIs: 1,000-iteration bootstrap (external: subsampled n=10,000). † = best in row. AUROC = Area Under the ROC Curve; AUPRC = Area Under the Precision-Recall Curve; ECE = Expected Calibration Error (10 uniform-width bins); PPV = Positive Predictive Value; NPV = Negative Predictive Value. Calibration slope and intercept from logistic recalibration on logit-transformed predictions (slope=1.0, intercept=0 = perfect calibration). Classification metrics at Youden-index threshold (maximizing sensitivity + specificity − 1) on internal validation set. Generalization gap = internal AUROC − external AUROC. Panel C ablation: racial AUROC disparity = max pairwise AUROC gap across White, Black, Hispanic, Asian subgroups on internal test set.

Global XGBoost feature importance — with urine output, GCS total, and lactate ranked among the top predictors by mean |SHAP| value — is presented in **Supplementary Figure S1**. Missingness SHAP analyses identified urine output (max racial differential in mean |SHAP|=0.0075) and lactate (0.0062) as the features with the greatest differential missingness-driven model reliance across racial groups, though absolute magnitudes were small, consistent with the subtle effect sizes identified in Steps 2–3 (**Supplementary Figures S2–S3; Supplementary Table S7**).

### 3.4 Model Performance on Internal and External Validation

All three models achieved strong internal discrimination, with XGBoost best across all dimensions (**Table 2**; **Supplementary Figure S5**). XGBoost achieved AUROC=0.910 (95% CI: 0.900–0.920; AUPRC=0.624; Brier=0.087; sensitivity=0.836; specificity=0.828). Random forest showed the lowest uncalibrated ECE (0.105) but a calibration slope of 1.417, indicating systematic risk underestimation. Logistic regression showed the largest ECE (0.220) and lowest calibration slope (0.871).

External validation revealed AUROC generalization gaps of 0.111–0.121 across all models (XGBoost: 0.799; 95% CI: 0.794–0.803). KS testing across 41 comparable predictors showed 39 with statistically significant distributional differences (p<0.001). The largest differences were GCS total (KS=0.699; MIMIC-IV mean=12.4 vs eICU mean=7.9) and Elixhauser comorbidity count (100% missing in eICU; **Supplementary Table S5**).

### 3.5 Formal Fairness Assessment

Full fairness metrics are presented in **Table 3** and **Figure 3**. Four key patterns emerged: racial TPR disparities were the largest fairness gap; calibration worsened substantially on external validation; ICU type showed clinically meaningful variation; and sex-based gaps were small across all criteria.

**Figure 3.**
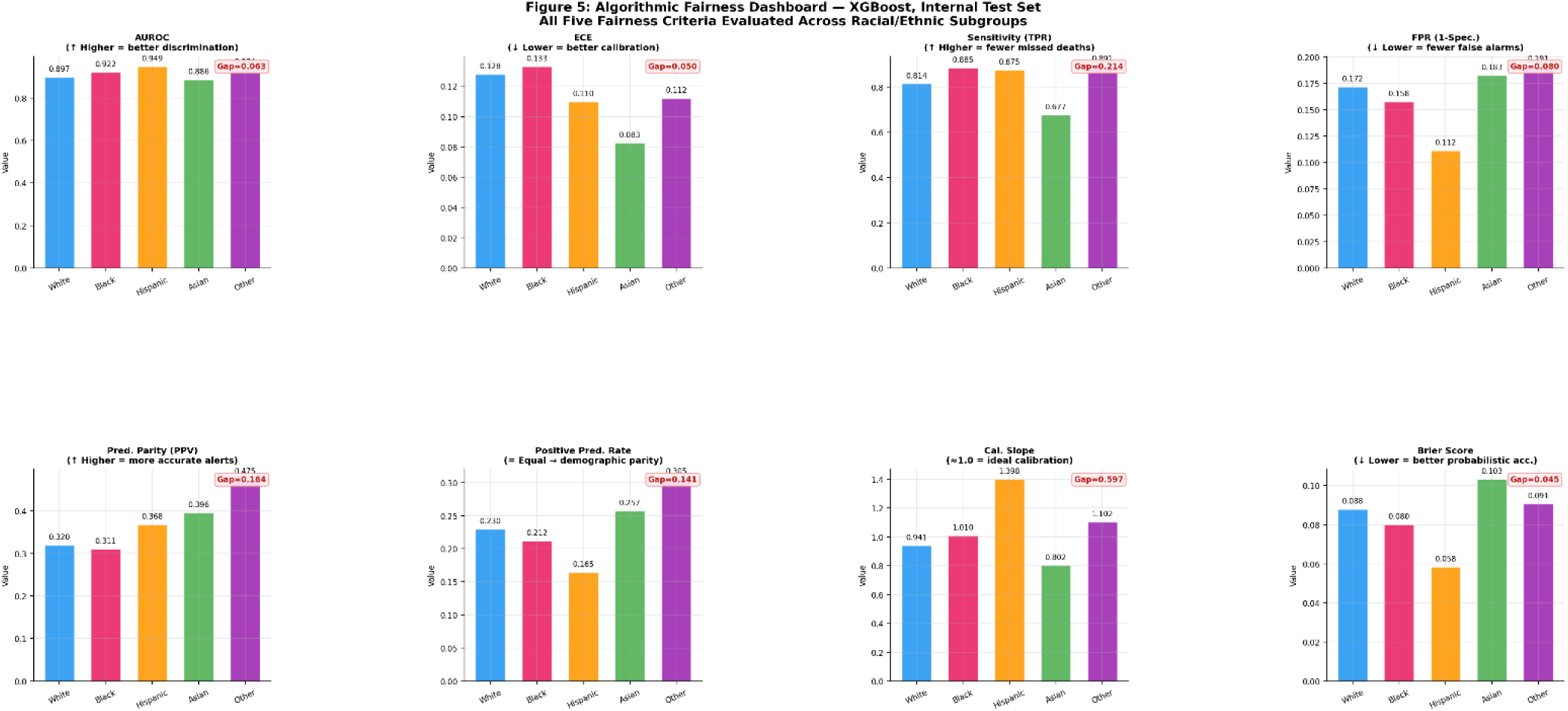
Fairness Dashboard — XGBoost, Racial and Ethnic Subgroups. Eight fairness metrics for XGBoost on the internal test set stratified by racial/ethnic group. Maximum between-group gap annotated per panel (red). Equalized-odds TPR gap (0.214) is the largest disparity. Subgroups with n<30 excluded. Internal test set; n=7,625.

**Table 3:**
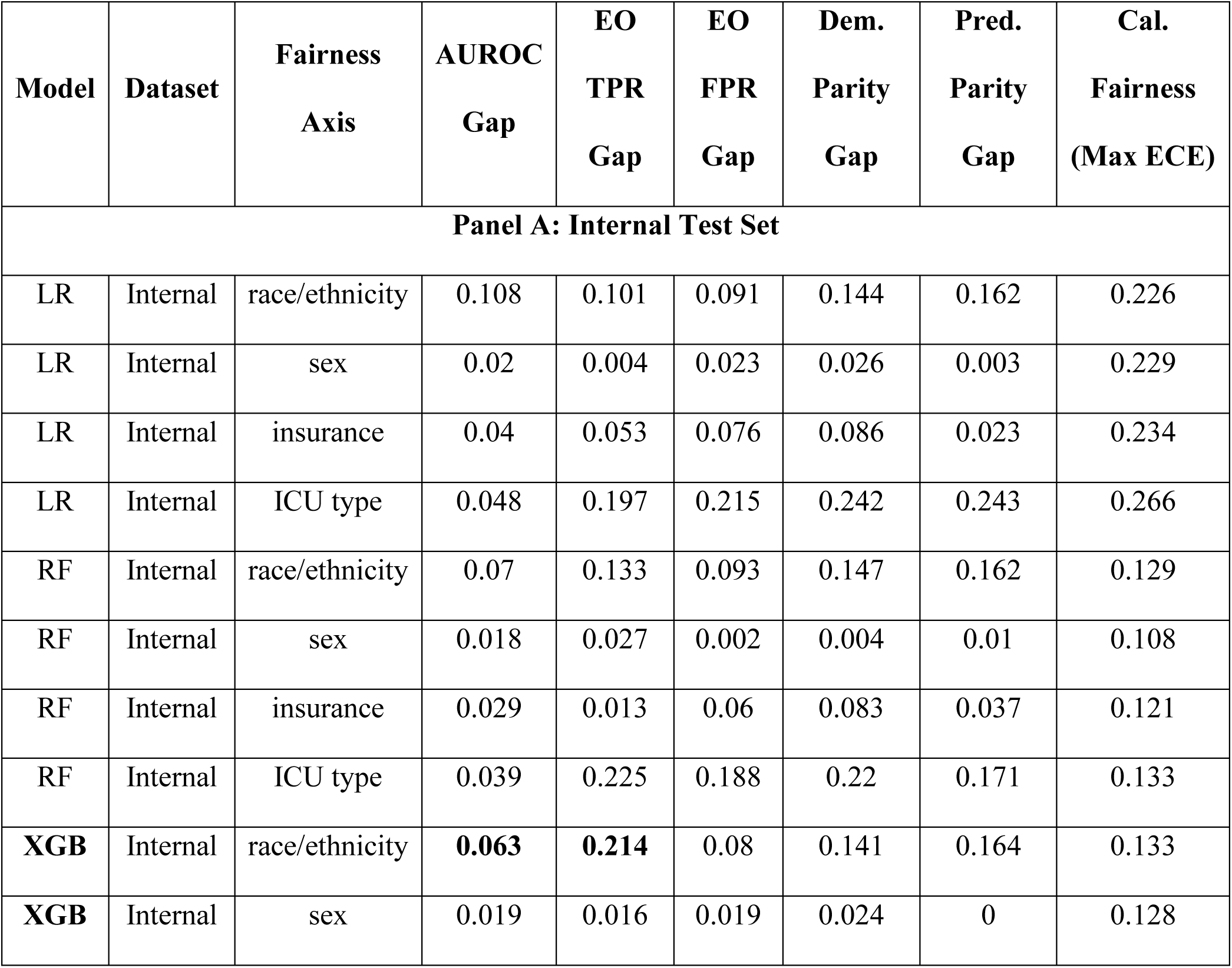
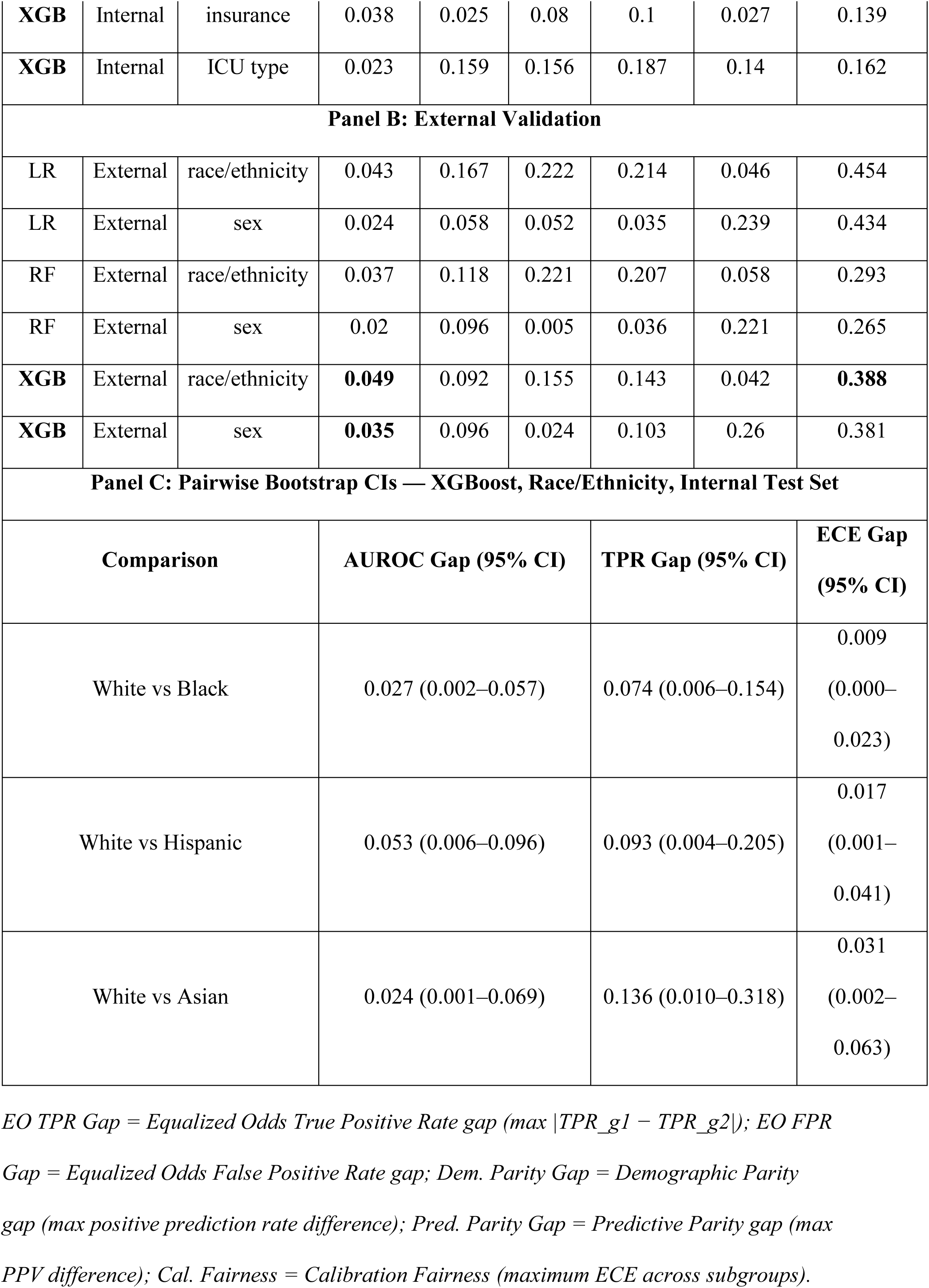
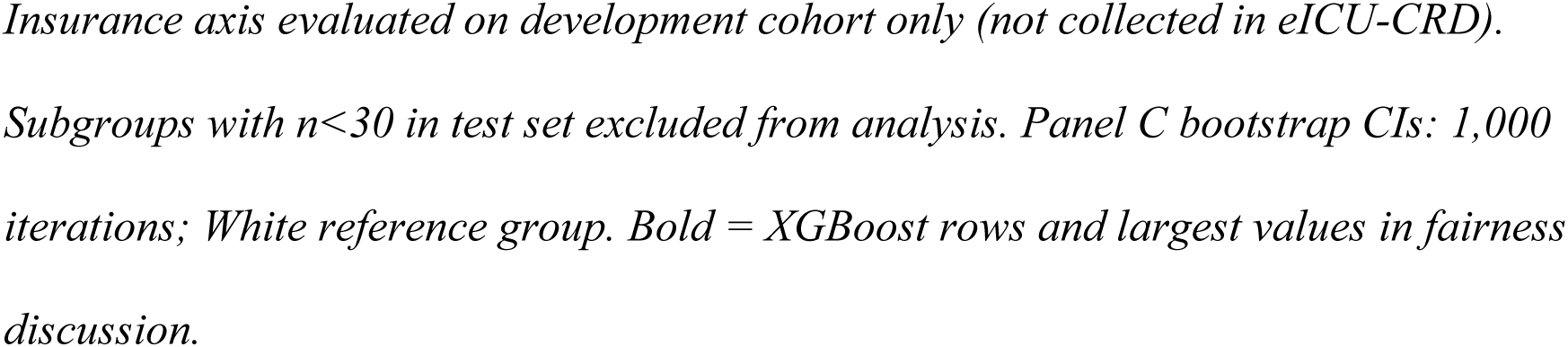
Formal Fairness Metrics — All Models, All Protected Attribute Axes. EO TPR Gap = Equalized Odds True Positive Rate gap (max |TPR_g1 − TPR_g2|); EO FPR Gap = Equalized Odds False Positive Rate gap; Dem. Parity Gap = Demographic Parity gap (max positive prediction rate difference); Pred. Parity Gap = Predictive Parity gap (max PPV difference); Cal. Fairness = Calibration Fairness (maximum ECE across subgroups). Insurance axis evaluated on development cohort only (not collected in eICU-CRD). Subgroups with n<30 in test set excluded from analysis. Panel C bootstrap CIs: 1,000 iterations; White reference group. Bold = XGBoost rows and largest values in fairness discussion.

On the internal test set, XGBoost showed a racial AUROC gap of 0.063, equalized-odds TPR gap of 0.214, and maximum-group ECE of 0.133. Pairwise bootstrap CIs confirmed significant White–Hispanic gaps (AUROC: 0.053, 95% CI: 0.006–0.096; TPR: 0.093, 95% CI: 0.004–0.205) and White–Asian TPR gap (0.136, 95% CI: 0.010–0.318); White–Black AUROC CIs partially overlapped zero (0.027, 95% CI: 0.002–0.057). ICU type showed a TPR gap of 0.159; sex gaps were ≤0.022 AUROC.

On external validation, racial discrimination disparities attenuated (AUROC gap=0.049) but calibration worsened markedly (MaxECE=0.388). Differing baseline mortality rates (7.8%–14.2% internally) rendered simultaneous satisfaction of equalized odds, predictive parity, and calibration fairness mathematically incompatible under Chouldechova’s impossibility theorem.^39^

### 3.6 Post-Hoc Recalibration

Recalibration results are in **Table 4** and the Pareto frontier in **Figure 4**. The central finding is that a single global Platt scaling step achieved 94% ECE reduction with no AUROC loss, transferred to external validation without retraining, and was Pareto-dominant on both dimensions.

**Figure 4.**
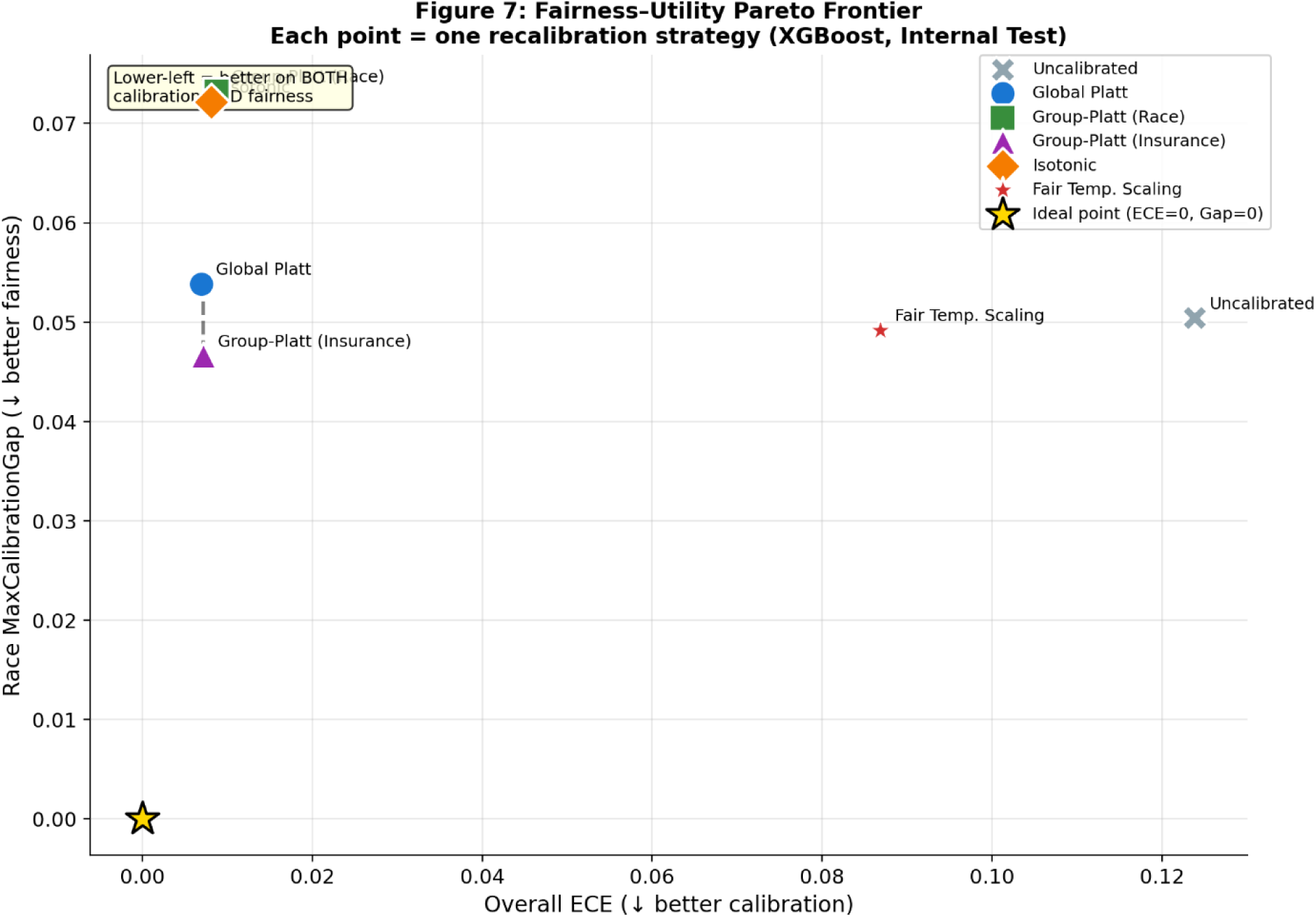
Fairness-Utility Pareto Frontier — Recalibration Strategies. Overall ECE (x-axis) versus racial calibration gap (y-axis) for six recalibration strategies applied to XGBoost on the internal test set. Lower-left = better on both dimensions. Dashed line = Pareto frontier. Gold star = ideal point. Global Platt and Group-Conditional Platt (Insurance) are Pareto-dominant.

**Table 4:** Post-Hoc Recalibration Strategy Comparison — XGBoost. All calibrators fitted on internal validation set only; no eICU data used. ECE = Expected Calibration Error (10 uniform bins). Race/Ins. Cal. Gap = max pairwise ECE across subgroups. Bold = best in column. † Group-Platt (Race) underperformed Global Platt due to small minority subgroup sizes (Hispanic n≈400, Asian n≈200). ‡ Slope <1.0 indicates logit-spread distortion when miscalibration is intercept-driven. Panel B: insurance strategies not applied to eICU.

| Strategy | Global ECE | Cal. Slope | Cal. Intercept | Global AUROC | Brier Score | Max Race ECE | Race Cal. Gap | Max Ins. ECE | Ins. Cal. Gap |
| --- | --- | --- | --- | --- | --- | --- | --- | --- | --- |
| **Panel A: Internal Test Set (n=7,625) — XGBoost** |  |  |  |  |  |  |  |  |  |
| **Uncalibrated** | 0.124 | 0.99 | −1.563 | 0.91 | 0.087 | 0.133 | 0.05 | 0.139 | 0.027 |
| Global Platt | **0.007** | 0.97 | −0.015 | 0.91 | 0.058 | 0.066 | 0.054 | 0.021 | 0.013 |
| Group-Platt (Race)<sup>†</sup> | 0.009 | 0.96 | −0.040 | 0.907 | 0.058 | 0.085 | 0.073 | 0.02 | 0.014 |
| Group-Platt (Insurance) | **0.007** | 0.97 | −0.021 | 0.91 | 0.058 | 0.058 | **0.047** | 0.018 | **0.007** |
| Isotonic<sup>‡</sup> | 0.008 | 0.84 | −0.220 | 0.909 | 0.059 | 0.083 | 0.072 | 0.024 | 0.016 |

| Fair Temp.<br>Scaling<sup>†‡</sup> | 0.087 | 0.48 | −1.563 | 0.91 | 0.092 | 0.117 | 0.049 | 0.099 | 0.025 |
| --- | --- | --- | --- | --- | --- | --- | --- | --- | --- |
| **Panel B: External Validation (n=137,773) — XGBoost** |  |  |  |  |  |  |  |  |  |
| Strategy | Global<br>ECE | Cal.<br>Slope | Cal.<br>Intercept | Global<br>AUROC | Brier<br>Score | Max<br>Race<br>ECE | Race<br>Cal.<br>Gap | *Insurance: not*<br>*available in*<br>*eICU* |  |
| Uncalibrated | 0.377 | 0.95 | −2.738 | 0.799 | 0.245 | 0.388 | 0.089 | — |  |
| Global Platt | 0.118 | 0.94 | −1.245 | 0.799 | 0.092 | 0.127 | 0.06 | — |  |
| Isotonic<sup>‡</sup> | **0.112** | 0.75 | −1.387 | 0.798 | 0.089 | 0.122 | **0.056** | — |  |
| Fair Temp.<br>Scaling<sup>†‡</sup> | 0.369 | 0.46 | −2.738 | 0.799 | 0.289 | 0.379 | 0.118 | — |  |

Internally, Global Platt reduced ECE from 0.124 to 0.007 (calibration slope=0.971; Brier: 0.087→0.058). Isotonic regression achieved the lowest ECE (0.008) but introduced slope distortion (0.835). Fairness-aware temperature scaling provided minimal improvement (ECE=0.087) because XGBoost’s miscalibration is intercept-driven — temperature scaling adjusts spread only and cannot correct intercept shifts.

Counterintuitively, Group-Conditional Platt by race produced a worse racial calibration gap (0.073) than Global Platt (0.054), attributed to overfitting in small minority validation subgroups (Hispanic n≈400, Asian n≈200; **Supplementary Table S6**). Group-Conditional Platt by insurance achieved the best racial gap (0.047). Subgroup calibration curves are shown in **Supplementary Figure S6**.

On external validation, Global Platt scaling reduced ECE from 0.377 to 0.118 (69% reduction) and isotonic regression achieved ECE=0.112, without any retraining on eICU data. Fairness-aware temperature scaling showed negligible external ECE improvement (0.370), consistent with its failure to address intercept-level miscalibration. The Pareto frontier (**Figure 4**) identified Global Platt and Group-Conditional Platt by insurance as the strategies closest to the ideal point (ECE=0, racial calibration gap=0), with Global Platt dominating on overall calibration and Group-Conditional Insurance dominating on racial calibration gap.

Group-stratified decision curve analysis confirmed consistent net benefit for XGBoost over treat-all and treat-none baselines across racial and insurance subgroups at clinically relevant threshold probabilities (**Supplementary Figure S4**).

## 4. DISCUSSION

### 4.1 Principal Findings

This study introduces MDLA and applies it within a comprehensive fairness audit of ICU mortality prediction across 188,600 patient-stays from two independent critical care databases. Three findings stand out. First, missingness patterns predicted racial group membership (AUROC=0.543; 95% CI: 0.540–0.546) and insurance type (AUROC=0.534; 0.531–0.539), with 18 of 43 features Bonferroni-significant — confirming that clinical measurement absence carries statistically significant demographic information. Second, models show evidence of relying on this signal: adding missingness indicators increased racial AUROC disparity by 10.7% without improving global performance, consistent with measurement absence constituting a latent bias pathway. Third, Global Platt scaling reduced ECE by 94% with zero AUROC loss, establishing post-hoc recalibration as a deployable fairness remedy. These results extend work on clinical AI fairness [48,49] and systematic ICU fairness auditing [50–52].

### 4.2 Missingness as a Bias Pathway

Prior work by Obermeyer et al [9]. demonstrated that biased proxy selection embeds racial disparities into clinical algorithms. Our results suggest an analogous but distinct mechanism: differential measurement practices carry demographic information within the data-generation process itself. Arterial blood gas measurements showed 14-percentage-point racial missingness gaps, consistent with known disparities in invasive monitoring access [53,54] and broader structural inequities in care [55]. All Cramér’s V values fell below 0.10 — the signal is subtle, which is precisely why a structured audit framework is needed.

XGBoost discrimination was competitive with published ICU mortality models [48,49]. The 0.111 generalization gap reflects structural cohort differences consistent with multi-site challenges documented across clinical AI [56–58], particularly GCS documentation practice variation (KS=0.699) and complete absence of Elixhauser comorbidity data in eICU.

### 4.3 Fairness Assessment and the Impossibility Theorem

The equalized-odds TPR gap of 0.214 substantially exceeds the AUROC gap of 0.063, consistent with Chouldechova’s impossibility theorem [39]: differing baseline mortality rates (7.8%–14.2%) make simultaneous satisfaction of equalized odds, predictive parity, and calibration fairness mathematically impossible. Institutions must therefore explicitly choose which criterion to prioritize [59,60]. For absolute risk prediction guiding clinical decisions, calibration fairness is the clinically appropriate primary criterion [15,16].

### 4.4 Recalibration Strategy Selection

Group-Conditional Platt by race produced a worse racial calibration gap (0.073) than Global Platt (0.054) — a counterintuitive finding attributable to small minority validation subgroups (Hispanic n≈400, Asian n≈200), where group-specific calibrators overfit and increase between-group ECE variance. This suggests a minimum subgroup size requirement not yet characterized in the fairness recalibration literature [46]. Fairness-aware temperature scaling also failed to improve calibration because XGBoost’s miscalibration is intercept-driven rather than spread-driven [45] — a distinction that should inform method selection in future deployments [61–63].

### 4.5 Limitations

Five limitations apply. First, both databases are US-based; missingness disparities may not generalize to health systems with different care or documentation structures [64,65]. Second, MDLA leakage AUROCs are modest (0.543 and 0.534) with all Cramér’s V below 0.10, indicating a subtle rather than severe pathway that warrants population-scale study. Third, all analyses are retrospective; prospective evaluation of clinical impact is needed [62]. Fourth, eICU lacks insurance and ICD data, limiting external fairness analyses. Fifth, the minimum calibration subgroup size finding (n≈200) requires formal simulation validation.

## 5. CONCLUSION

Clinical measurement absence is not simply a data quality problem — it is a structured demographic signal that standard fairness audits do not evaluate. Through MDLA, this study provides a reproducible four-step framework for detecting this upstream bias pathway across 188,600 ICU patient-stays from two institutionally diverse databases. Missingness patterns alone predicted racial group membership above chance, and ablation analyses indicated model reliance on this signal — increasing racial AUROC disparity by 10.7% when missingness indicators were added. A five-criteria fairness audit revealed a 0.214 equalized-odds TPR gap irresolvable through model tuning, given mathematical incompatibility across differing subgroup base rates. Global Platt recalibration reduced calibration error by 94% without AUROC loss and transferred to external validation without retraining. These findings are associative rather than causal and prospective validation remains needed — but they make a specific, actionable case: missingness-aware auditing and calibration-aware evaluation should be standard components of clinical AI validation before deployment.

## ACKNOWLEDGEMENTS

The authors gratefully acknowledge the patients whose de-identified clinical data, contributed through Beth Israel Deaconess Medical Center and the participating institutions of the eICU Collaborative Research Database, made this research possible. We thank the PhysioNet platform and the MIMIC and eICU research teams for maintaining open access to these critical care databases and for the credentialing infrastructure that enables reproducible research.

The computational analyses in this study were conducted using publicly available open-source software, including Python, scikit-learn, XGBoost, SHAP, and statsmodels, whose developer communities we acknowledge.

No external funding was received for this research. The views expressed are solely those of the authors.

## CREDIT AUTHOR CONTRIBUTIONS

### Krutarth Patel

Conceptualization; Methodology; Software; Formal Analysis; Investigation; Data Curation; Validation; Visualization; Writing – Original Draft; Writing – Review and Editing; Project Administration.

### Phanindra Beedala

Data Curation; Writing – Original Draft.

## ETHICAL STATEMENT

This study used exclusively de-identified, secondary patient data obtained from two publicly available critical care databases — MIMIC-IV v2.2 and the eICU Collaborative Research Database v2.0 — both accessible through PhysioNet under credentialed data use agreements (https://physionet.org). All data were de-identified prior to public release by the originating institutions in accordance with the Health Insurance Portability and Accountability Act (HIPAA) Safe Harbor method. No patient contact occurred, no identifiable information was accessed or retained, and no new data collection was performed. Accordingly, this study was exempt from institutional review board (IRB) oversight under 45 CFR §46.104(d)(4), which covers research involving the use of existing, publicly available de-identified data. The authors completed the required PhysioNet credentialing and data use agreement training prior to data access.

## DATA AVAILABILITY STATEMENT

The clinical data used in this study are publicly available through PhysioNet (https://physionet.org) subject to credentialed data use agreement registration and completion of the required human research training:

- **MIMIC-IV v2.2** (development cohort): Johnson, A., Bulgarelli, L., Pollard, T., Horng, S., Celi, L.A., & Mark, R. (2023). MIMIC-IV (version 2.2). PhysioNet. https://doi.org/10.13026/6mm1-ek67
- **eICU Collaborative Research Database v2.0** (external validation cohort): Pollard, T., Johnson, A., Raffa, J., Celi, L.A., Mark, R., & Badawi, O. (2018). eICU Collaborative Research Database (version 2.0). PhysioNet. https://doi.org/10.13026/C2WM1R

The complete analysis code — including all data preprocessing pipelines, MDLA implementation, model training, fairness evaluation, recalibration strategies, and figure generation — is publicly available at [<u>mdla-icu-fairness-audit</u>]. The repository reproduces all tables and figures reported in this manuscript from the cached intermediate outputs, which are also provided to avoid reprocessing the full raw databases. All random seeds are fixed (seed=42) to ensure exact reproducibility of results.

Derived aggregate outputs (Tables 1–4, Figures 1–4, and all supplementary tables) are included directly in the manuscript and its supplementary materials and do not contain patient-level information.

## PATIENT AND PUBLIC INVOLVEMENT

No patients or members of the public were involved in the design, conduct, or reporting of this study. The research used retrospective, de-identified electronic health record data from publicly available critical care databases.

## DECLARATION OF GENERATIVE AI AND AI-ASSISTED TECHNOLOGIES IN THE MANUSCRIPT PREPARATION PROCESS

During the preparation of this work the authors used Claude (Anthropic), Gemini (Google) and ChatGPT (OpenAI) in order to assist with language editing, grammatical refinement, and manuscript organization, and used ChatGPT (OpenAI) in order to support code debugging and troubleshooting during model development and analysis. After using these tools/services, the authors reviewed and edited the content as needed and take full responsibility for the content of the published article.

---

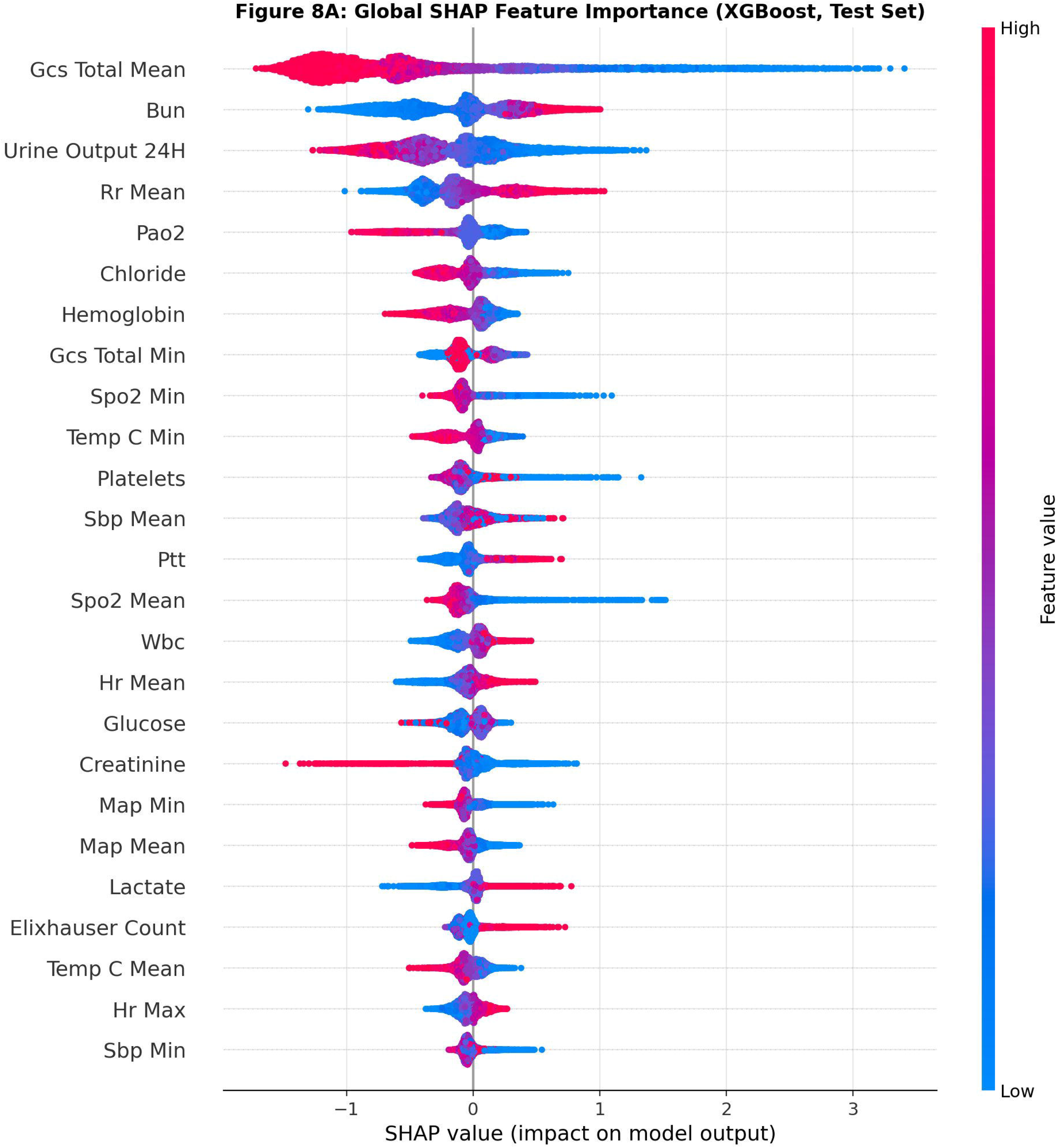

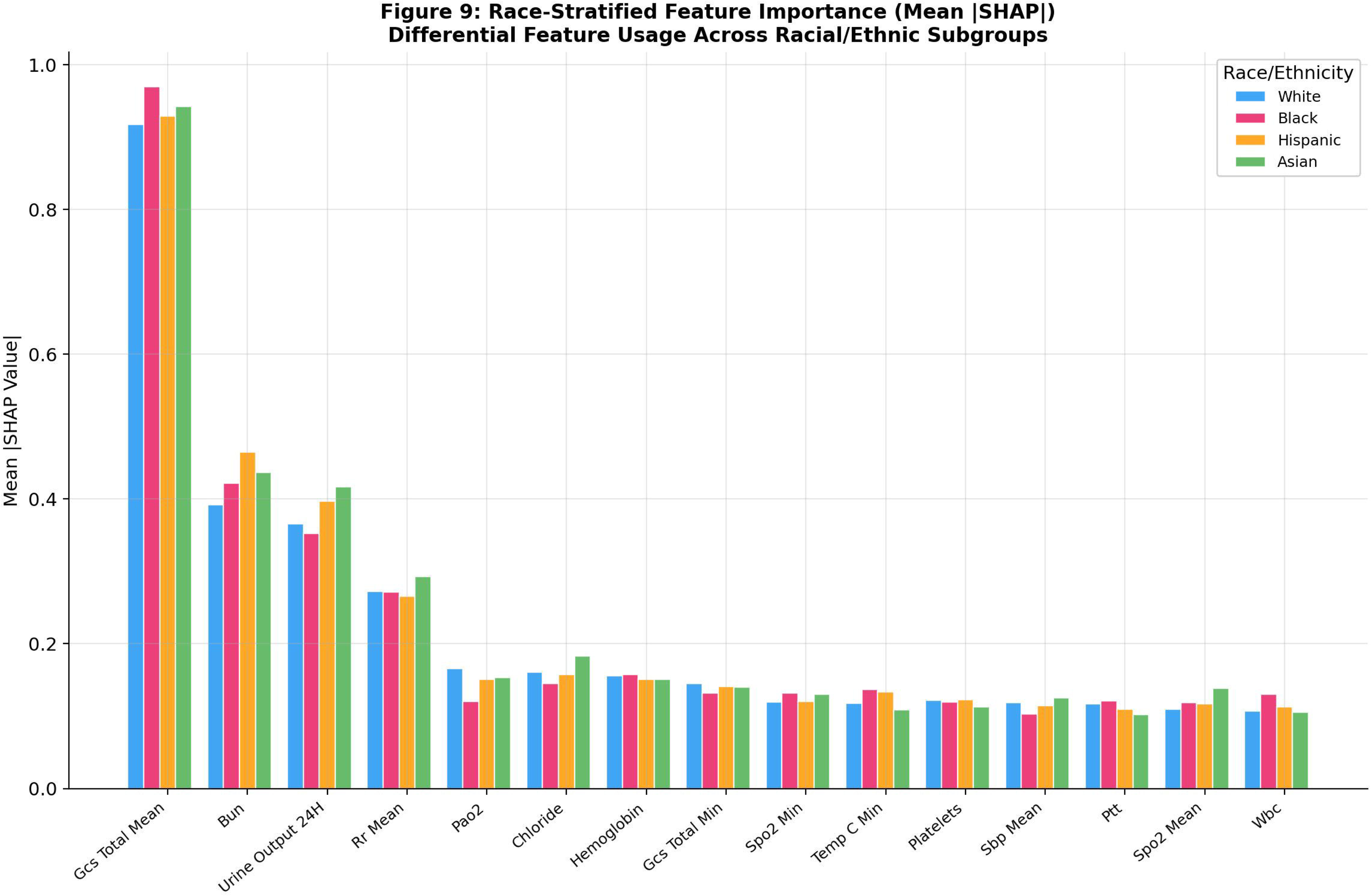

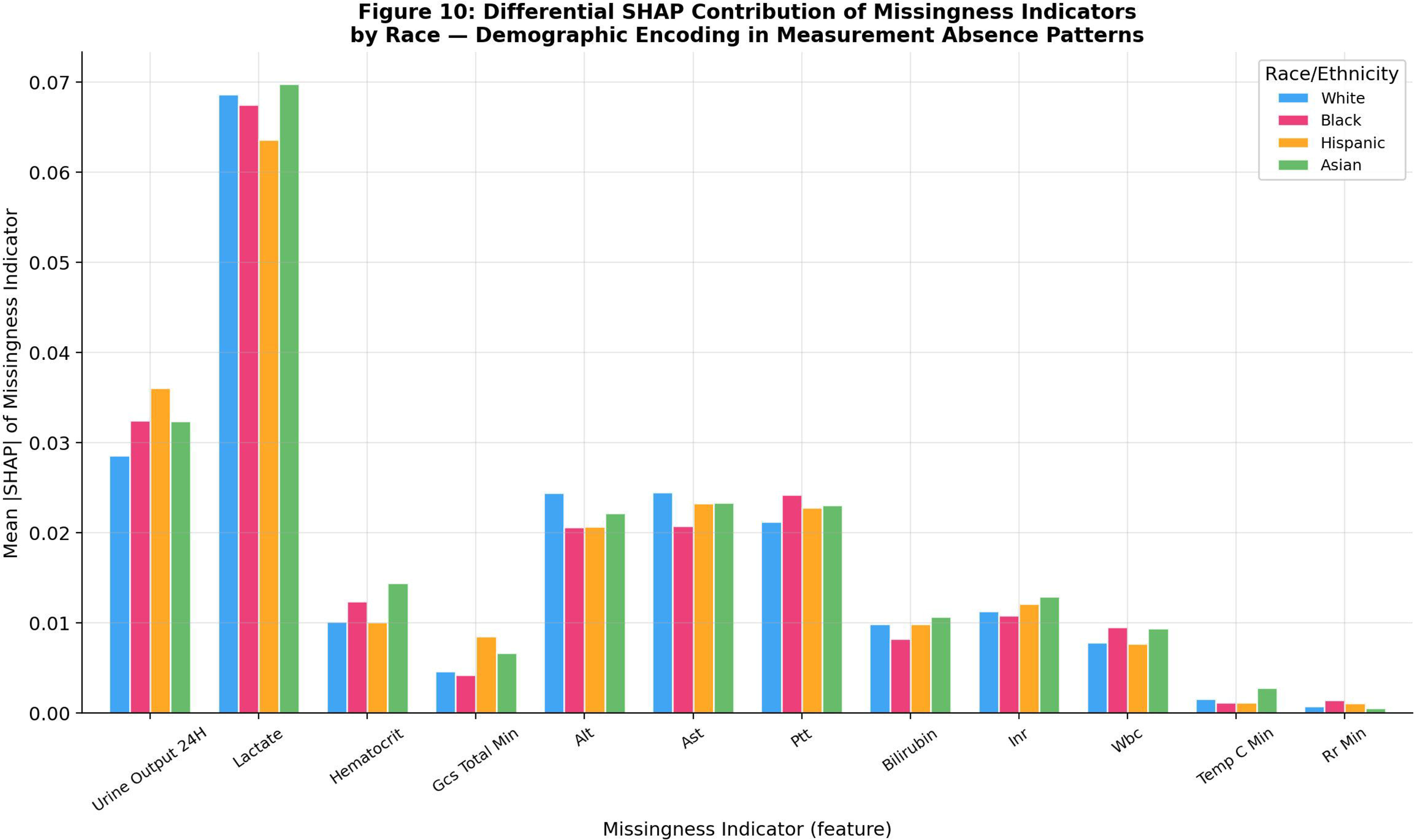

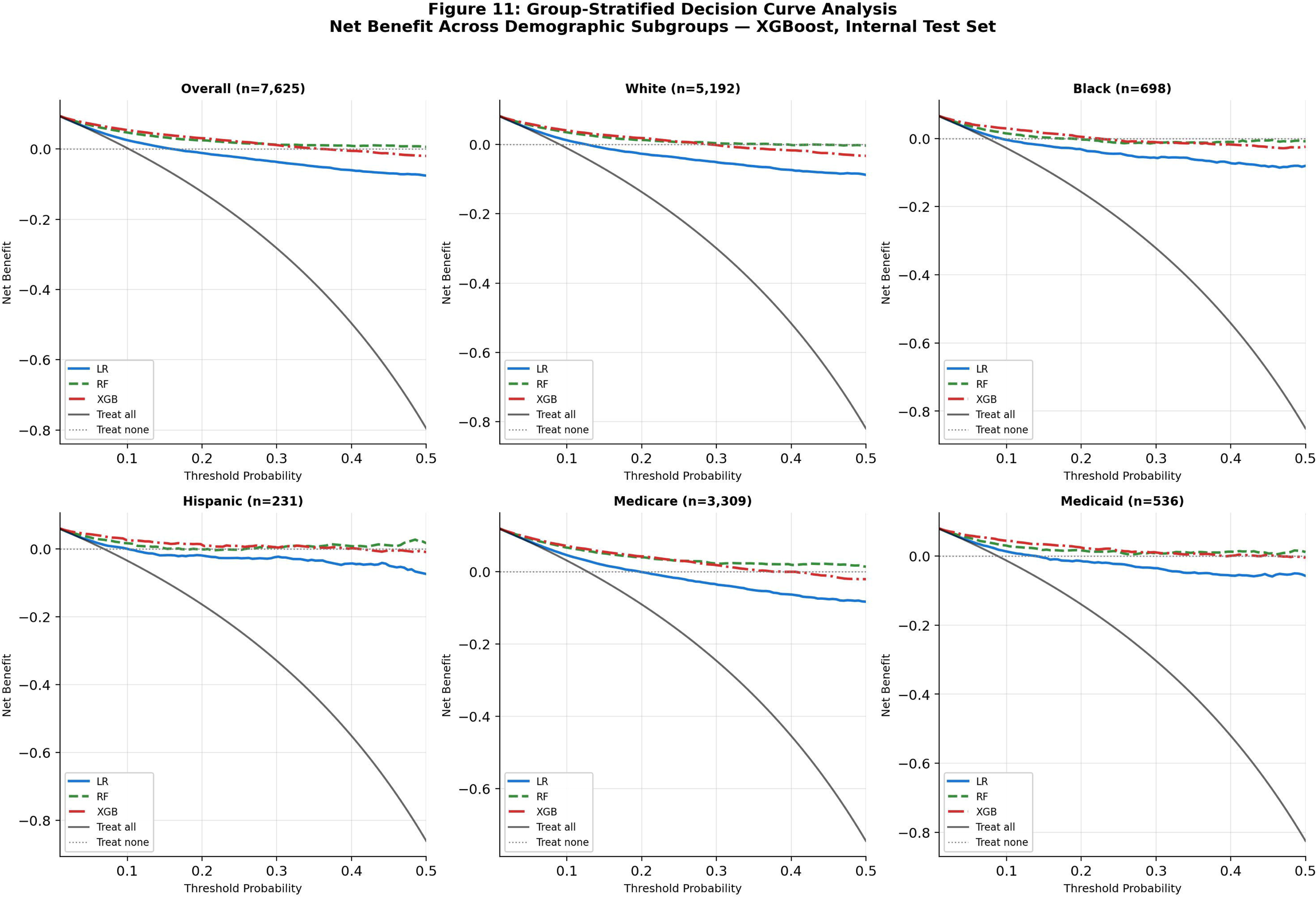

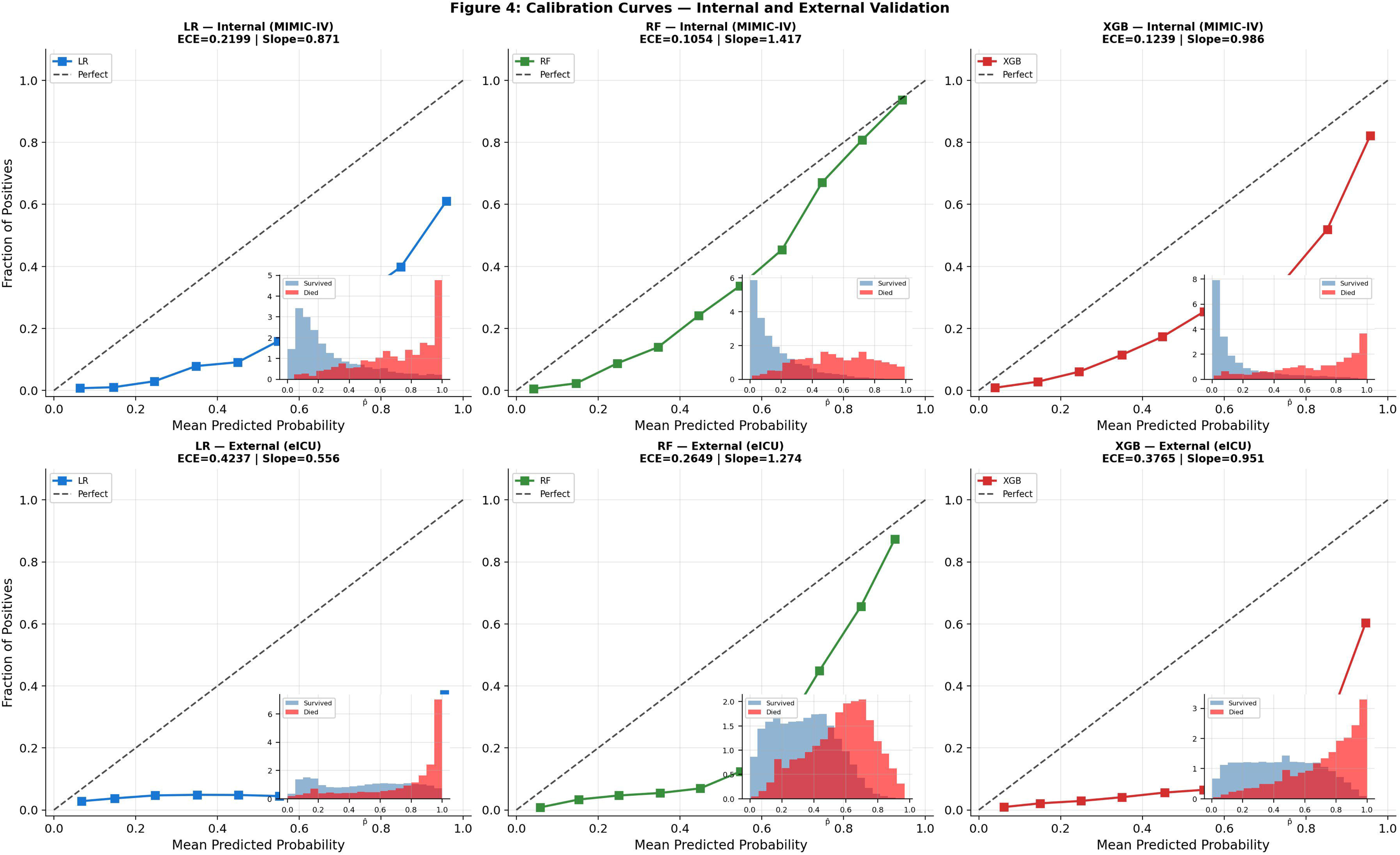

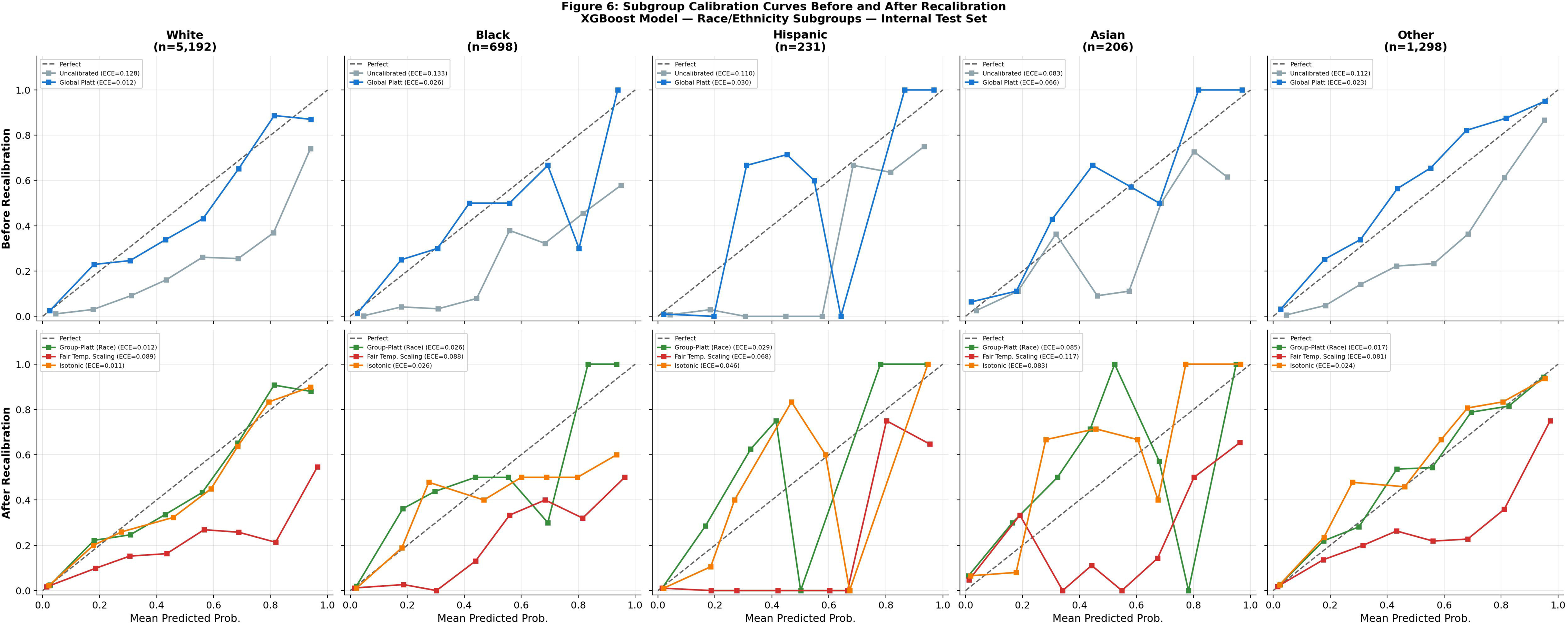

**Table S1.** Hyperparameter Grid Search Specifications and Selected Values for All Three Classifier Models.

| Parameter | Grid Values Searched | Selected Value | Notes |
| --- | --- | --- | --- |
| **Logistic Regression** (L2, lbfgs solver) |  |  | 5-fold CV AUROC = 0.8830 |
| Regularization strength (C) | {0.01, 0.1, 1.0, 10.0} | **10** |  |
| Penalty | {l2} | l2 | Fixed |
| Solver | {lbfgs} | lbfgs | Fixed |
| Class weights | {balanced} | balanced |  |
| Max iterations | {2000} | 2000 |  |
| Random state | {42} | 42 |  |
| **Random Forest** |  |  | 5-fold CV AUROC = 0.9023 |
| n_estimators | {200, 500} | **500** |  |
| max_depth | {None, 10, 20} | **None (unlimited)** |  |
| min_samples_leaf | {10, 20} | **10** |  |
| max_features | {sqrt} | sqrt | Fixed |
| Class weights | {balanced} | balanced | Fixed |
| Random state | {42} | 42 | Fixed |
| **XGBoost** |  |  | 5-fold CV AUROC = 0.9086 |
| n_estimators | {300, 500} | **300** |  |
| max_depth | {4, 6} | **6** |  |
| learning_rate | {0.05, 0.1} | **0.05** |  |
| min_child_weight | {5, 10} | **10** |  |
| subsample | {0.8} | 0.8 | Fixed |
| colsample_bytree | {0.8} | 0.8 | Fixed |
| scale_pos_weight | {neg/pos ratio} | **8.8** | Derived from training class imbalance |
| eval_metric | {logloss} | logloss | Fixed |
| Random state | {42} | 42 | Fixed |

*Hyperparameter search space, selected values, and cross-validation performance for Logistic Regression, Random Forest, and XGBoost. Grid search was performed using 5-fold stratified cross-validation on the training set (n = 35,578), optimizing for AUROC. All models used a fixed random state of 42 for reproducibility.*

**Table S2.** Missingness Rates by Feature and Demographic Group.

| **Feature** | **Overall Miss. Rate** | **White Miss. Rate** | **Black Miss. Rate** | **Hispanic Miss. Rate** | **Asian Miss. Rate** | **Race Max Gap** | **Medicare Miss.** | **Medicaid Miss.** | **Other/Unknown Miss.** | **Insurance Max Gap** |
| --- | --- | --- | --- | --- | --- | --- | --- | --- | --- | --- |
| **Paco2** | 0.4642 | 0.4787 | 0.5165 | 0.4830 | 0.4804 | 0.1404 | 0.4622 | 0.4638 | 0.4659 | 0.0037 |
| **Pao2** | 0.4641 | 0.4786 | 0.5165 | 0.4830 | 0.4804 | 0.1404 | 0.4621 | 0.4638 | 0.4658 | 0.0037 |
| **Ph** | 0.4422 | 0.4559 | 0.4943 | 0.4593 | 0.4554 | 0.1364 | 0.4410 | 0.4385 | 0.4437 | 0.0052 |
| **Ptt** | 0.1931 | 0.1911 | 0.2722 | 0.2123 | 0.2115 | 0.1184 | 0.1861 | 0.2155 | 0.1960 | 0.0295 |
| **Lactate** | 0.4643 | 0.4765 | 0.5092 | 0.4807 | 0.4713 | 0.1184 | 0.4590 | 0.4558 | 0.4700 | 0.0142 |
| **Inr** | 0.1867 | 0.1851 | 0.2607 | 0.2123 | 0.2039 | 0.1135 | 0.1784 | 0.2073 | 0.1909 | 0.0288 |
| **Alt** | 0.6004 | 0.6222 | 0.5790 | 0.5718 | 0.5937 | 0.0875 | 0.6050 | 0.5048 | 0.6100 | 0.1052 |
| **Ast** | 0.5992 | 0.6209 | 0.5775 | 0.5695 | 0.5914 | 0.0861 | 0.6033 | 0.5048 | 0.6092 | 0.1043 |
| **Bilirubin** | 0.6042 | 0.6252 | 0.5790 | 0.5765 | 0.5929 | 0.0809 | 0.6073 | 0.5136 | 0.6144 | 0.1008 |
| **Albumin** | 0.7403 | 0.7562 | 0.7358 | 0.7115 | 0.7341 | 0.0674 | 0.7458 | 0.6697 | 0.7457 | 0.0761 |
| **Urine Output 24H** | 0.0305 | 0.0277 | 0.0569 | 0.0364 | 0.0302 | 0.0305 | 0.0381 | 0.0261 | 0.0246 | 0.0135 |
| **Temp C Max** | 0.0326 | 0.0327 | 0.0183 | 0.0277 | 0.0287 | 0.0227 | 0.0343 | 0.0226 | 0.0326 | 0.0117 |
| **Temp C Mean** | 0.0326 | 0.0327 | 0.0183 | 0.0277 | 0.0287 | 0.0227 | 0.0343 | 0.0226 | 0.0326 | 0.0117 |
| **Temp C Min** | 0.0326 | 0.0327 | 0.0183 | 0.0277 | 0.0287 | 0.0227 | 0.0343 | 0.0226 | 0.0326 | 0.0117 |
| **Platelets** | 0.0347 | 0.0327 | 0.0451 | 0.0404 | 0.0415 | 0.0124 | 0.0323 | 0.0380 | 0.0363 | 0.0057 |
| **Hemoglobin** | 0.0353 | 0.0333 | 0.0451 | 0.0398 | 0.0446 | 0.0118 | 0.0328 | 0.0391 | 0.0369 | 0.0062 |
| **Wbc** | 0.0354 | 0.0335 | 0.0451 | 0.0387 | 0.0431 | 0.0116 | 0.0328 | 0.0391 | 0.0371 | 0.0062 |
| **Hematocrit** | 0.0320 | 0.0301 | 0.0412 | 0.0346 | 0.0400 | 0.0111 | 0.0300 | 0.0361 | 0.0331 | 0.0060 |
| **Bun** | 0.0298 | 0.0285 | 0.0321 | 0.0375 | 0.0347 | 0.0090 | 0.0279 | 0.0339 | 0.0308 | 0.0059 |
| **Creatinine** | 0.0295 | 0.0282 | 0.0321 | 0.0364 | 0.0347 | 0.0082 | 0.0277 | 0.0333 | 0.0306 | 0.0057 |
| **Bicarbonate** | 0.0298 | 0.0283 | 0.0319 | 0.0364 | 0.0347 | 0.0081 | 0.0281 | 0.0336 | 0.0307 | 0.0055 |
| **Potassium** | 0.0296 | 0.0284 | 0.0313 | 0.0364 | 0.0347 | 0.0080 | 0.0276 | 0.0322 | 0.0310 | 0.0046 |
| **Chloride** | 0.0290 | 0.0277 | 0.0315 | 0.0352 | 0.0340 | 0.0075 | 0.0271 | 0.0322 | 0.0302 | 0.0051 |
| **Sodium** | 0.0291 | 0.0278 | 0.0308 | 0.0352 | 0.0340 | 0.0074 | 0.0273 | 0.0325 | 0.0301 | 0.0052 |
| **Glucose** | 0.0340 | 0.0333 | 0.0336 | 0.0392 | 0.0355 | 0.0060 | 0.0316 | 0.0344 | 0.0361 | 0.0045 |
| **Gcs Total Min** | 0.0046 | 0.0045 | 0.0035 | 0.0081 | 0.0060 | 0.0046 | 0.0051 | 0.0061 | 0.0041 | 0.0020 |

| Feature | Overall Miss. Rate | White Miss. Rate | Black Miss. Rate | Hispanic Miss. Rate | Asian Miss. Rate | Race Max Gap | Medicare Miss. | Medicaid Miss. | Other/Unknown Miss. | Insurance Max Gap |
| --- | --- | --- | --- | --- | --- | --- | --- | --- | --- | --- |
| Gcs Total Mean | 0.0046 | 0.0045 | 0.0035 | 0.0081 | 0.0060 | 0.0046 | 0.0051 | 0.0061 | 0.0041 | 0.0020 |
| Sbp Mean | 0.0041 | 0.0042 | 0.0041 | 0.0058 | 0.0015 | 0.0043 | 0.0041 | 0.0047 | 0.0039 | 0.000746 |
| Sbp Max | 0.0041 | 0.0042 | 0.0041 | 0.0058 | 0.0015 | 0.0043 | 0.0041 | 0.0047 | 0.0039 | 0.000746 |
| Sbp Min | 0.0041 | 0.0042 | 0.0041 | 0.0058 | 0.0015 | 0.0043 | 0.0041 | 0.0047 | 0.0039 | 0.000746 |
| Map Mean | 0.0018 | 0.0021 | 0.0015 | 0.0035 | 0.0023 | 0.0029 | 0.0017 | 0.0028 | 0.0018 | 0.0010 |
| Map Min | 0.0018 | 0.0021 | 0.0015 | 0.0035 | 0.0023 | 0.0029 | 0.0017 | 0.0028 | 0.0018 | 0.0010 |
| Map Max | 0.0018 | 0.0021 | 0.0015 | 0.0035 | 0.0023 | 0.0029 | 0.0017 | 0.0028 | 0.0018 | 0.0010 |
| Spo2 Mean | 0.0021 | 0.0021 | 0.0026 | 0.0035 | 0.000755 | 0.0027 | 0.0021 | 0.0039 | 0.0018 | 0.0020 |
| Spo2 Max | 0.0021 | 0.0021 | 0.0026 | 0.0035 | 0.000755 | 0.0027 | 0.0021 | 0.0039 | 0.0018 | 0.0020 |
| Spo2 Min | 0.0021 | 0.0021 | 0.0026 | 0.0035 | 0.000755 | 0.0027 | 0.0021 | 0.0039 | 0.0018 | 0.0020 |
| Hr Min | 0.0013 | 0.0015 | 0.0013 | 0.0029 | 0.000755 | 0.0026 | 0.0013 | 0.0022 | 0.0012 | 0.0010 |
| Hr Max | 0.0013 | 0.0015 | 0.0013 | 0.0029 | 0.000755 | 0.0026 | 0.0013 | 0.0022 | 0.0012 | 0.0010 |
| Hr Mean | 0.0013 | 0.0015 | 0.0013 | 0.0029 | 0.000755 | 0.0026 | 0.0013 | 0.0022 | 0.0012 | 0.0010 |
| Rr Mean | 0.0030 | 0.0035 | 0.0026 | 0.0035 | 0.0015 | 0.0020 | 0.0029 | 0.0036 | 0.0031 | 0.000685 |
| Rr Max | 0.0030 | 0.0035 | 0.0026 | 0.0035 | 0.0015 | 0.0020 | 0.0029 | 0.0036 | 0.0031 | 0.000685 |
| Rr Min | 0.0030 | 0.0035 | 0.0026 | 0.0035 | 0.0015 | 0.0020 | 0.0029 | 0.0036 | 0.0031 | 0.000685 |
| Troponin | 1.0000 | 1.0000 | 1.0000 | 1.0000 | 1.0000 | 0.0000 | 1.0000 | 1.0000 | 1.0000 | 0.0000 |
| Elixhauser Count | 0.0000 | 0.0000 | 0.0000 | 0.0000 | 0.0000 | 0.0000 | 0.0000 | 0.0000 | 0.0000 | 0.0000 |

Values represent proportion of missing observations (0.000–1.000) per feature per demographic group. Race Max Gap = maximum absolute difference in missingness rate across racial/ethnic groups. Insurance Max Gap = maximum absolute difference across insurance categories. Development cohort; n=50,827.

**Table S3.** Chi-Square Tests and Cramér’s V Effect Sizes — Race-Missingness Associations (All 43 Features)

| feature | chi2 | p_value | cramers_v | dof | p_bonferroni | significant |
| --- | --- | --- | --- | --- | --- | --- |
| lab_paco2 | 364.3087 | 1.4262e-77 | 0.0847 | 4.0000 | 5.9901e-76 | True |
| lab_pao2 | 363.8505 | 1.7912e-77 | 0.0846 | 4.0000 | 7.5230e-76 | True |
| lab_ph | 339.2814 | 3.6146e-72 | 0.0817 | 4.0000 | 1.5181e-70 | True |
| lab_ptt | 283.2999 | 4.3298e-60 | 0.0747 | 4.0000 | 1.8185e-58 | True |
| lab_inr | 270.5565 | 2.4201e-57 | 0.0730 | 4.0000 | 1.0165e-55 | True |
| lab_lactate | 255.5475 | 4.1533e-54 | 0.0709 | 4.0000 | 1.7444e-52 | True |
| lab_alt | 244.4801 | 1.0059e-51 | 0.0694 | 4.0000 | 4.2250e-50 | True |
| lab_ast | 238.2846 | 2.1720e-50 | 0.0685 | 4.0000 | 9.1223e-49 | True |
| lab_bilirubin | 216.6842 | 9.6919e-46 | 0.0653 | 4.0000 | 4.0706e-44 | True |
| lab_albumin | 177.2756 | 2.8681e-37 | 0.0591 | 4.0000 | 1.2046e-35 | True |
| urine_output_24h | 126.0212 | 2.7612e-26 | 0.0498 | 4.0000 | 1.1597e-24 | True |
| temp_c_min | 52.0093 | 0.000000 | 0.0320 | 4.0000 | 0.000000 | True |
| temp_c_max | 52.0093 | 0.000000 | 0.0320 | 4.0000 | 0.000000 | True |
| temp_c_mean | 52.0093 | 0.000000 | 0.0320 | 4.0000 | 0.000000 | True |
| lab_platelets | 22.6524 | 0.000149 | 0.0211 | 4.0000 | 0.0062 | True |
| lab_hemoglobin | 21.5003 | 0.000252 | 0.0206 | 4.0000 | 0.0106 | True |
| lab_hematocrit | 20.1603 | 0.000464 | 0.0199 | 4.0000 | 0.0195 | True |
| lab_wbc | 19.0932 | 0.000753 | 0.0194 | 4.0000 | 0.0316 | True |
| map_mean | 12.3933 | 0.0147 | 0.0156 | 4.0000 | 0.6155 | False |
| map_max | 12.3933 | 0.0147 | 0.0156 | 4.0000 | 0.6155 | False |
| map_min | 12.3933 | 0.0147 | 0.0156 | 4.0000 | 0.6155 | False |
| hr_min | 11.2799 | 0.0236 | 0.0149 | 4.0000 | 0.9909 | False |
| hr_max | 11.2799 | 0.0236 | 0.0149 | 4.0000 | 0.9909 | False |
| hr_mean | 11.2799 | 0.0236 | 0.0149 | 4.0000 | 0.9909 | False |
| lab_bicarbonate | 9.0489 | 0.0599 | 0.0133 | 4.0000 | 1.0000 | False |
| rr_min | 8.3896 | 0.0783 | 0.0128 | 4.0000 | 1.0000 | False |
| rr_max | 8.3896 | 0.0783 | 0.0128 | 4.0000 | 1.0000 | False |
| rr_mean | 8.3896 | 0.0783 | 0.0128 | 4.0000 | 1.0000 | False |
| lab_bun | 8.3425 | 0.0798 | 0.0128 | 4.0000 | 1.0000 | False |
| lab_creatinine | 8.3113 | 0.0808 | 0.0128 | 4.0000 | 1.0000 | False |
| lab_chloride | 7.7670 | 0.1005 | 0.0124 | 4.0000 | 1.0000 | False |
| lab_potassium | 6.9788 | 0.1370 | 0.0117 | 4.0000 | 1.0000 | False |
| lab_sodium | 6.8521 | 0.1439 | 0.0116 | 4.0000 | 1.0000 | False |
| gcs_total_min | 6.5985 | 0.1587 | 0.0114 | 4.0000 | 1.0000 | False |
| gcs_total_mean | 6.5985 | 0.1587 | 0.0114 | 4.0000 | 1.0000 | False |
| spo2_min | 4.0257 | 0.4025 | 0.0089 | 4.0000 | 1.0000 | False |
| spo2_mean | 4.0257 | 0.4025 | 0.0089 | 4.0000 | 1.0000 | False |
| spo2_max | 4.0257 | 0.4025 | 0.0089 | 4.0000 | 1.0000 | False |
| sbp_mean | 3.6338 | 0.4578 | 0.0085 | 4.0000 | 1.0000 | False |
| sbp_max | 3.6338 | 0.4578 | 0.0085 | 4.0000 | 1.0000 | False |
| sbp_min | 3.6338 | 0.4578 | 0.0085 | 4.0000 | 1.0000 | False |
| lab_glucose | 2.9465 | 0.5668 | 0.0076 | 4.0000 | 1.0000 | False |

Chi-square tests of independence between each binary missingness indicator and racial group membership. Bonferroni correction applied across 43 simultaneous tests ( $\alpha=0.05/43=0.00116$ ). dof = degrees of freedom. Cramér's V: <0.10 = small effect, 0.10–0.30 = medium, >0.30 = large. All 18 significant associations have $V<0.10$ . miss\_troponin excluded (constant=1 for all training observations).

**Table S4.**
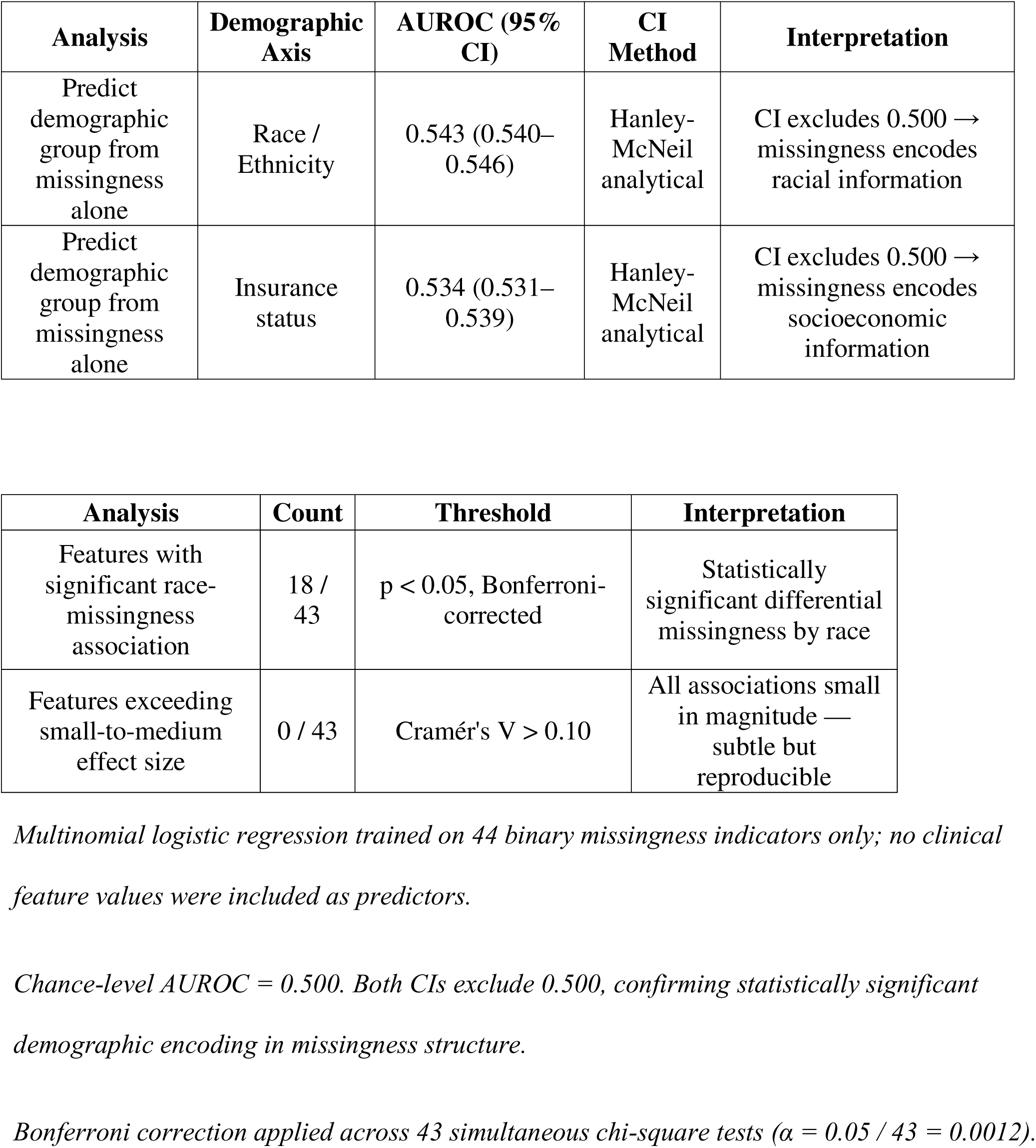
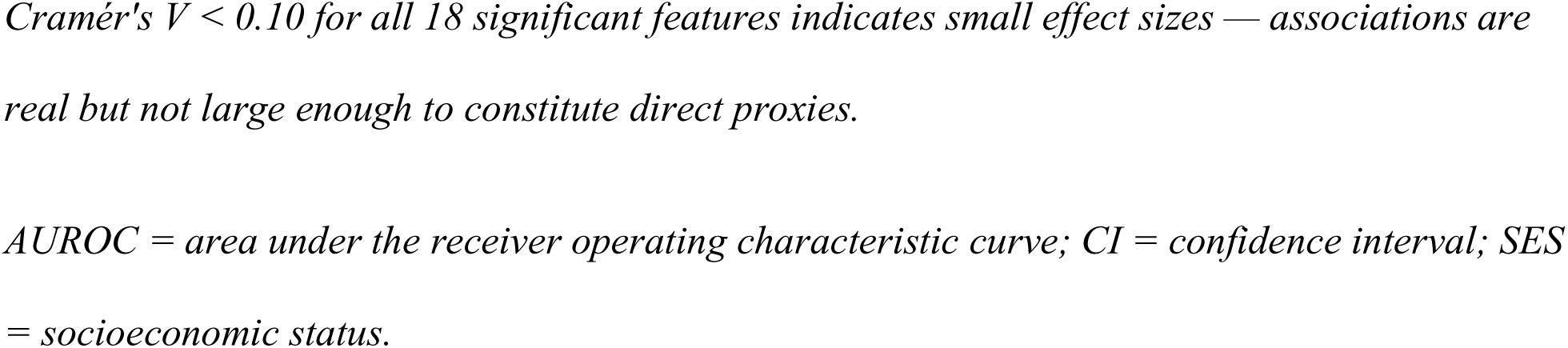
Demographic Predictability from Missingness Patterns Alone — AUROC, Bootstrap 95% Confidence Intervals, and Feature-Level Association Summary.

**Table S5.** Kolmogorov-Smirnov Distributional Shift Analysis — MIMIC-IV vs. eICU-CRD (41 Predictors)

| Feature | KS Stat | p_value | MIMIC Mean | eICU Mean | MIMIC Miss% | eICU Miss% | Significant |
| --- | --- | --- | --- | --- | --- | --- | --- |
| gcs_total_mean | 0.6990 | 0.0000 | 12.4182 | 7.9210 | 0.0046 | 0.2757 | True |
| gcs_total_min | 0.5772 | 0.0000 | 10.3960 | 4.8335 | 0.0046 | 0.2757 | True |
| temp_c_max | 0.3478 | 0.0000 | 37.3346 | 38.8885 | 0.0326 | 0.9063 | True |
| temp_c_min | 0.3379 | 0.0000 | 36.3154 | 35.6335 | 0.0326 | 0.9063 | True |
| spo2_min | 0.3036 | 0.0000 | 91.9842 | 86.6416 | 0.0021 | 0.0401 | True |
| temp_c_mean | 0.2759 | 0.0000 | 36.8304 | 38.0016 | 0.0326 | 0.9063 | True |
| sbp_max | 0.2542 | 0.0000 | 147.9395 | 166.5031 | 0.0041 | 0.7612 | True |
| map_max | 0.2523 | 0.0000 | 103.8526 | 139.4942 | 0.0018 | 0.7596 | True |
| sbp_min | 0.2358 | 0.0000 | 93.3569 | 82.1238 | 0.0041 | 0.7612 | True |
| lab_pao2 | 0.1908 | 0.0000 | 177.0743 | 137.8144 | 0.4641 | 0.6870 | True |
| rr_max | 0.1791 | 0.0000 | 27.4959 | 32.7378 | 0.0030 | 0.1001 | True |
| rr_min | 0.1789 | 0.0000 | 12.3432 | 10.3100 | 0.0030 | 0.1001 | True |
| map_min | 0.1461 | 0.0000 | 60.5251 | 52.2094 | 0.0018 | 0.7596 | True |
| lab_albumin | 0.1390 | 9.8456e-182 | 3.1753 | 2.9342 | 0.7403 | 0.5792 | True |
| lab_ptt | 0.1337 | 2.1214e-282 | 37.1414 | 40.0504 | 0.1931 | 0.7652 | True |
| lab_ast | 0.1122 | 5.5706e-163 | 211.2610 | 157.4506 | 0.5992 | 0.6061 | True |
| lab_ph | 0.1066 | 1.7649e-168 | 7.3645 | 7.3620 | 0.4422 | 0.6923 | True |
| hr_max | 0.1031 | 0.0000 | 102.4038 | 107.3146 | 0.0013 | 0.0305 | True |
| lab_bicarbonate | 0.1015 | 1.0554e-307 | 22.9672 | 23.8007 | 0.0298 | 0.1880 | True |
| lab_creatinine | 0.0925 | 5.7300e-260 | 1.3058 | 1.4582 | 0.0295 | 0.1361 | True |
| lab_bilirubin | 0.0884 | 1.7872e-99 | 1.8503 | 1.1478 | 0.6042 | 0.6161 | True |
| lab_hematocrit | 0.0874 | 1.6571e-230 | 32.7528 | 33.9712 | 0.0320 | 0.1548 | True |
| lab_hemoglobin | 0.0780 | 9.8631e-183 | 10.8583 | 11.2531 | 0.0353 | 0.1602 | True |
| lab_inr | 0.0695 | 1.8016e-92 | 1.4477 | 1.5738 | 0.1867 | 0.6618 | True |
| lab_lactate | 0.0694 | 6.3473e-63 | 2.3711 | 2.6778 | 0.4643 | 0.7601 | True |
| lab_alt | 0.0628 | 4.9858e-51 | 150.0594 | 99.6545 | 0.6004 | 0.6108 | True |
| sbp_mean | 0.0610 | 4.8030e-65 | 119.0266 | 120.5888 | 0.0041 | 0.7612 | True |
| rr_mean | 0.0608 | 3.7854e-116 | 18.9782 | 19.4292 | 0.0030 | 0.1001 | True |
| lab_paco2 | 0.0579 | 5.7226e-49 | 42.1806 | 42.7672 | 0.4642 | 0.6901 | True |
| hr_min | 0.0522 | 1.6789e-87 | 69.9166 | 67.5096 | 0.0013 | 0.0305 | True |
| lab_potassium | 0.0506 | 2.5083e-78 | 4.1748 | 4.1001 | 0.0296 | 0.1307 | True |
| map_mean | 0.0467 | 2.0905e-38 | 78.9395 | 80.5592 | 0.0018 | 0.7596 | True |
| spo2_max | 0.0382 | 5.3216e-47 | 99.4165 | 99.4982 | 0.0021 | 0.0401 | True |
| lab_glucose | 0.0375 | 7.0914e-43 | 141.1452 | 146.8098 | 0.0340 | 0.1420 | True |
| spo2_mean | 0.0374 | 4.2878e-45 | 96.8355 | 96.6060 | 0.0021 | 0.0401 | True |
| lab_bun | 0.0370 | 7.3206e-42 | 23.9193 | 24.9664 | 0.0298 | 0.1397 | True |
| lab_sodium | 0.0362 | 3.2163e-40 | 138.5136 | 138.2895 | 0.0291 | 0.1357 | True |
| hr_mean | 0.0309 | 6.5082e-31 | 84.2282 | 83.9647 | 0.0013 | 0.0305 | True |
| lab_platelets | 0.0211 | 1.1192e-13 | 205.1714 | 202.9702 | 0.0347 | 0.1794 | True |
| lab_wbc | 0.0092 | 0.0058 | 11.9759 | 12.1346 | 0.0354 | 0.1697 | False |
| lab_chloride | 0.0084 | 0.0140 | 104.5329 | 104.5197 | 0.0290 | 0.1420 | False |

*Two-sample KS tests between MIMIC-IV v2.2 (development) and eICU-CRD v2.0 (external validation) distributions for 41 comparable predictors. Elixhauser comorbidity count and 24-hour urine output excluded (100% missing in eICU). p-values reported without multiple testing correction; analysis is exploratory. MIMIC Miss% and eICU Miss% = proportion of missing observations per cohort. Significant = $p < 0.001$ .*

**Table S6.**
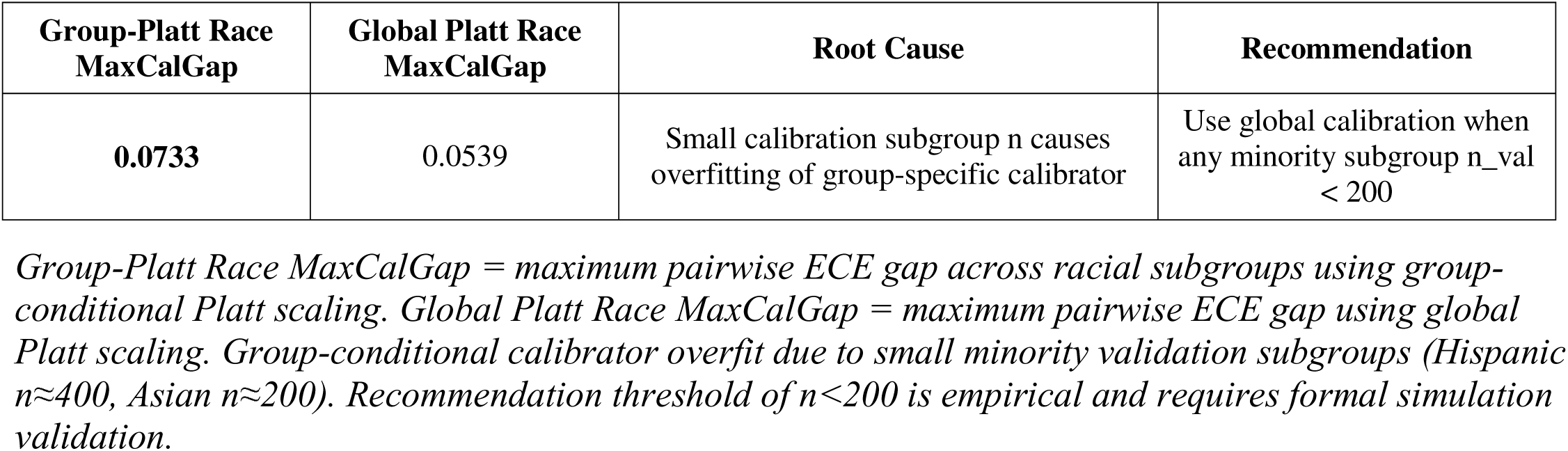
Group-Conditional vs. Global Platt Scaling — Calibration Gap Comparison and Root Cause.

| Group-Platt Race MaxCalGap | Global Platt Race MaxCalGap | Root Cause | Recommendation |
| --- | --- | --- | --- |
| 0.0733 | 0.0539 | Small calibration subgroup n causes overfitting of group-specific calibrator | Use global calibration when any minority subgroup n_val < 200 |

*Group-Platt Race MaxCalGap = maximum pairwise ECE gap across racial subgroups using group-conditional Platt scaling. Global Platt Race MaxCalGap = maximum pairwise ECE gap using global Platt scaling. Group-conditional calibrator overfit due to small minority validation subgroups (Hispanic $n \approx 400$ , Asian $n \approx 200$ ). Recommendation threshold of $n < 200$ is empirical and requires formal simulation validation.*

**Table S7.** Race-Stratified Mean |SHAP| Values per Feature — XGBoost, Internal Test Set.

|  | **White** | **Black** | **Hispanic** | **Asian** | **max_diff** |
| --- | --- | --- | --- | --- | --- |
| **Hr Min** | 0.0000 | 0.0000 | 0.0000 | 0.0000 | 0.0000 |
| **Hr Max** | 0.0000 | 0.0000 | 0.0000 | 0.0000 | 0.0000 |
| **Hr Mean** | 0.0000 | 0.0000 | 0.0000 | 0.0000 | 0.0000 |
| **Sbp Min** | 0.0000 | 0.0000 | 0.0000 | 0.0000 | 0.0000 |
| **Sbp Max** | 0.0000 | 0.0000 | 0.0000 | 0.0000 | 0.0000 |
| **Sbp Mean** | 0.0000 | 0.0000 | 0.0000 | 0.0000 | 0.0000 |
| **Map Min** | 0.0000 | 0.0000 | 0.0000 | 0.0000 | 0.0000 |
| **Map Max** | 0.0000 | 0.0000 | 0.0000 | 0.0000 | 0.0000 |
| **Map Mean** | 0.0000 | 0.0000 | 0.0000 | 0.0000 | 0.0000 |
| **Rr Min** | 0.000781 | 0.0014 | 0.0011 | 0.000591 | 0.000836 |
| **Rr Max** | 0.000236 | 0.000411 | 0.000286 | 0.000202 | 0.000209 |
| **Rr Mean** | 0.0000 | 0.0000 | 0.0000 | 0.0000 | 0.0000 |
| **Spo2 Min** | 0.0000 | 0.0000 | 0.0000 | 0.0000 | 0.0000 |
| **Spo2 Max** | 0.0000 | 0.0000 | 0.0000 | 0.0000 | 0.0000 |
| **Spo2 Mean** | 0.0000 | 0.0000 | 0.0000 | 0.0000 | 0.0000 |
| **Temp C Min** | 0.0016 | 0.0012 | 0.0012 | 0.0028 | 0.0016 |
| **Temp C Max** | 0.0000 | 0.0000 | 0.0000 | 0.0000 | 0.0000 |
| **Temp C Mean** | 0.0000 | 0.0000 | 0.0000 | 0.0000 | 0.0000 |
| **Gcs Total Min** | 0.0046 | 0.0042 | 0.0085 | 0.0067 | 0.0043 |
| **Gcs Total Mean** | 0.0000 | 0.0000 | 0.0000 | 0.0000 | 0.0000 |
| **Creatinine** | 0.0018 | 0.0019 | 0.0020 | 0.0027 | 0.000809 |
| **Bun** | 0.0000 | 0.0000 | 0.0000 | 0.0000 | 0.0000 |
| **Lactate** | 0.0686 | 0.0675 | 0.0636 | 0.0698 | 0.0062 |
| **Hemoglobin** | 0.000650 | 0.000811 | 0.000728 | 0.000677 | 0.000161 |
| **Hematocrit** | 0.0102 | 0.0124 | 0.0101 | 0.0145 | 0.0044 |
| **Platelets** | 0.000186 | 0.000239 | 0.000190 | 0.000212 | 0.000054 |
| **Wbc** | 0.0078 | 0.0095 | 0.0077 | 0.0094 | 0.0019 |
| **Sodium** | 0.000230 | 0.000209 | 0.000160 | 0.000397 | 0.000237 |
| **Potassium** | 0.000696 | 0.000786 | 0.0010 | 0.000596 | 0.000405 |
| **Bicarbonate** | 0.000507 | 0.000478 | 0.000653 | 0.000459 | 0.000195 |
| **Chloride** | 0.0000 | 0.0000 | 0.0000 | 0.0000 | 0.0000 |
| **Glucose** | 0.000973 | 0.000935 | 0.0014 | 0.000982 | 0.000442 |
| **Pao2** | 0.0048 | 0.0052 | 0.0046 | 0.0045 | 0.000637 |
| **Paco2** | 0.0044 | 0.0047 | 0.0044 | 0.0040 | 0.000653 |
| **Ph** | 0.0062 | 0.0060 | 0.0067 | 0.0067 | 0.000690 |
| **Alt** | 0.0244 | 0.0207 | 0.0207 | 0.0222 | 0.0038 |
| **Ast** | 0.0245 | 0.0208 | 0.0233 | 0.0233 | 0.0038 |
| **Bilirubin** | 0.0099 | 0.0083 | 0.0099 | 0.0107 | 0.0024 |
| **Albumin** | 0.0024 | 0.0026 | 0.0027 | 0.0027 | 0.000379 |
| **Inr** | 0.0113 | 0.0108 | 0.0121 | 0.0129 | 0.0021 |
| **Ptt** | 0.0212 | 0.0242 | 0.0228 | 0.0231 | 0.0030 |
| **Troponin** | 0.0000 | 0.0000 | 0.0000 | 0.0000 | 0.0000 |
| **Urine Output 24H** | 0.0286 | 0.0325 | 0.0361 | 0.0324 | 0.0075 |
| **Elixhauser Count** | 0.0000 | 0.0000 | 0.0000 | 0.0000 | 0.0000 |

**Table S8.** TRIPOD+AI Reporting Checklist.

| Item | Section | Description | Status |
| --- | --- | --- | --- |
| 1a | Title | Identifies the study as a fairness audit of a prediction model | ☐ |
| 1b | Abstract | Structured abstract with background, methods, fairness findings, conclusions | ☐ |
| 2 | Background | Fairness audit framed in context of algorithmic disparities in ICU care | ☐ |
| 3a | Source of data | MIMIC-IV v2.2 (development) + eICU v2.0 (external validation) | ☐ |
| 3b | Dates | MIMIC-IV: 2008–2022; eICU: 2014–2015 | ☐ |
| 4 | Participants | Eligibility criteria identical across datasets | ☐ |
| 5 | Outcome | In-hospital mortality, operationalized consistently | ☐ |
| 6 | Predictors | 43 first-24h features (20 vitals, 22 labs, 2 clinical); gcs_total and lab_troponin excluded due to 100% missingness — retained as binary missingness indicators per item 8a | ☐ |
| 7a | Sample size | No formal sample size calculation; >50K train, >100K validation | ☐ |
| 8a | Missing data | Binary missingness indicators; median imputation from training set | ☐ |
| 8b | Missingness analysis | Novel: demographic-structured missingness as bias pathway | ☐ |
| 10a | Model development | LR, RF, XGBoost; 5-fold CV; composite calibration criterion | ☐ |
| 10b | Calibration | ECE, slope, intercept; subgroup-specific; four recalibration strategies | ☐ |
| 10c | Performance | AUROC, AUPRC, Brier, sensitivity, specificity, PPV, NPV, F1 | ☐ |
| 12 | External validation | eICU: no retraining; preprocessing from MIMIC-IV only | ☐ |
| 13a | Fairness audit | Five formal fairness criteria: EO, EqOpp, DP, PP, CalFair | ☐ |
| 13b | Protected attributes | Race/ethnicity, sex, insurance, age group, ICU type | ☐ |
| 13c | Subgroup recalibration | Four strategies compared; Pareto frontier analysis | ☐ |
| 14 | Discrimination | SHAP (global + race-stratified + missingness SHAP) | ☐ |
| 15 | DCA | Group-stratified DCA across race and insurance subgroups | ☐ |
| 17 | Reporting guideline | TRIPOD+AI (Collins et al., BMJ 2024) | ☐ |
| 19 | Code availability | Full pipeline available at GitHub | ☐ |

*TRIPOD+AI = Transparent Reporting of a multivariable prediction model for Individual Prognosis or Diagnosis + Artificial Intelligence. Reference: Collins GS et al. BMJ 2024;385:e078378. = criterion met. All applicable items are satisfied.*

## REFERENCES

[1] Kannan S, Mahajan P, Harrington P, et al. Critical care utilization and outcomes in the United States, 2019–2022. Crit Care Med. 2024;52(3):e134–e143.

[2] Society of Critical Care Medicine. Critical Care Statistics. 2024. https://www.sccm.org/Communications/Critical-Care-Statistics. Accessed April 28, 2026.

[3] Halpern NA, Goldman DA, Tan KS, Pastores SM. Trends in critical care beds and use among population groups and Medicare and Medicaid beneficiaries in the United States: 2000–2010. Crit Care Med. 2016;44(8):1490–1499.

[4] Adhikari NK, Fowler RA, Bhagwanjee S, Rubenfeld GD. Critical care and the global burden of critical illness in adults. Lancet. 2010;376(9749):1339–1346.

[5] Johnson AEW, Ghassemi MM, Nemati S, et al. Machine learning and decision support in critical care. Proc IEEE. 2016;104(2):444–466.

[6] Thorsen-Meyer HC, Nielsen AB, Nielsen AP, Kaas-Hansen BS, Toft P, Schierbeck J, et al. Dynamic and explainable machine learning prediction of mortality in patients in the intensive care unit. Lancet Digit Health. 2020;2(4):e179–e191.

[7] Hyland SL, Faltys M, Hüser M, Lyu X, Gumbsch T, Esteban C, et al. Early prediction of circulatory failure in the intensive care unit using machine learning. Nat Med. 2020;26(3):364–373.

[8] Awad A, Bader-El-Den M, McNicholas J, Briggs J. Early hospital mortality prediction of intensive care unit patients using an ensemble learning approach. Int J Med Inform. 2017;108:185–195.

[9] Obermeyer Z, Powers B, Vogeli C, Mullainathan S. Dissecting racial bias in an algorithm used to manage the health of populations. Science. 2019;366(6464):447–453.

[10] Celi LA, Cellini J, Charpignon ML, Dee EC, Dernoncourt F, Eber R, et al. Sources of bias in artificial intelligence that perpetuate healthcare disparities — a global review. PLOS Digit Health. 2022;1(3):e0000022.

[11] Ashana DC, Anesi GL, Liu VX, et al. Equitably allocating resources during crises: racial differences in mortality prediction models. Am J Respir Crit Care Med. 2021;204(2):178–186.

[12] Seyyed-Kalantari L, Zhang H, McDermott MBA, Chen IY, Ghassemi M. Underdiagnosis bias of artificial intelligence algorithms applied to chest radiographs in under-served patient populations. Nat Med. 2021;27(12):2176–2182.

[13] Rajkomar A, Hardt M, Howell MD, Corrado G, Chin MH. Ensuring fairness in machine learning to advance health equity. Ann Intern Med. 2018;169(12):866–872.

[14] Chen IY, Pierson E, Rose S, Joshi S, Ferryman K, Ghassemi M. Ethical machine learning in healthcare. Annu Rev Biomed Data Sci. 2021;4:123–144.

[15] van Calster B, McLernon DJ, van Smeden M, et al. Calibration: the Achilles heel of predictive analytics. BMC Med. 2019;17(1):230.

[16] van Calster B, Nieboer D, Vergouwe Y, De Cock B, Pencina MJ, Steyerberg EW. A calibration hierarchy for risk models was defined from easy to use to subtle to measure. J Clin Epidemiol. 2016;74:167–176.

[17] Agniel D, Kohane IS, Weber GM. Biases in electronic health record data due to processes within the healthcare system: retrospective observational study. BMJ. 2018;361:k1479.

[18] Cismondi F, Celi LA, Fialho AS, et al. Reducing unnecessary lab testing in the ICU with artificial intelligence. Int J Med Inform. 2013;82(5):345–358.

[19] Little RJA, Rubin DB. Statistical Analysis with Missing Data. 3rd ed. Hoboken, NJ: Wiley; 2019.

[20] Saria S, Butte A, Sheikh A. Better medicine through machine learning: what’s real, and what’s artificial? PLOS Med. 2018;15(12):e1002721.

[21] Collins GS, Moons KGM, Dhiman P, Riley RD, Beam AL, Van Calster B, et al. TRIPOD+AI statement: updated guidance for reporting clinical prediction models that use regression or machine learning. BMJ. 2024;385:e078378.

[22] Moons KGM, Altman DG, Reitsma JB, et al. Transparent Reporting of a multivariable prediction model for Individual Prognosis or Diagnosis (TRIPOD): explanation and elaboration. Ann Intern Med. 2015;162(1):W1–W73.

[23] Pedregosa F, Varoquaux G, Gramfort A, Michel V, Thirion B, Grisel O, et al. Scikit-learn: machine learning in Python. J Mach Learn Res. 2011;12:2825–2830.

[24] Chen T, Guestrin C. XGBoost: a scalable tree boosting system. In: Proc 22nd ACM SIGKDD Int Conf Knowl Discov Data Min. 2016:785–794.

[25] Lundberg SM, Lee SI. A unified approach to interpreting model predictions. Adv Neural Inf Process Syst. 2017;30:4765–4774.

[26] Lundberg SM, Erion G, Chen H, et al. From local explanations to global understanding with explainable AI for trees. Nat Mach Intell. 2020;2(1):56–67.

[27] Johnson AEW, Bulgarelli L, Shen L, et al. MIMIC-IV, a freely accessible electronic health record dataset. Sci Data. 2023;10(1):1.

[28] Pollard TJ, Johnson AEW, Raffa JD, Celi LA, Mark RG, Badawi O. The eICU Collaborative Research Database, a freely available multi-center database for critical care research. Sci Data. 2018;5:180178.

[29] Goldberger AL, Amaral LAN, Glass L, Hausdorff JM, Ivanov PC, Mark RG, et al. PhysioBank, PhysioToolkit, and PhysioNet: components of a new research resource for complex physiologic signals. Circulation. 2000;101(23):e215–e220.

[30] Elixhauser A, Steiner C, Harris DR, Coffey RM. Comorbidity measures for use with administrative data. Med Care. 1998;36(1):8–27.

[31] van Walraven C, Austin PC, Jennings A, Quan H, Forster AJ. A modification of the Elixhauser comorbidity measures into a point system for hospital death using administrative data. Med Care. 2009;47(6):626–633.

[32] van Buuren S. Flexible Imputation of Missing Data. 2nd ed. Boca Raton: CRC Press; 2018.

[33] Hanley JA, McNeil BJ. The meaning and use of the area under a receiver operating characteristic (ROC) curve. Radiology. 1982;143(1):29–36.

[34] Breiman L. Random forests. Mach Learn. 2001;45(1):5–32.

[35] Steyerberg EW, Vickers AJ, Cook NR, Gerds T, Gonen M, Obuchowski N, et al. Assessing the performance of prediction models: a framework for traditional and novel measures. Epidemiology. 2010;21(1):128–138.

[36] DeLong ER, DeLong DM, Clarke-Pearson DL. Comparing the areas under two or more correlated receiver operating characteristic curves: a nonparametric approach. Biometrics. 1988;44(3):837–845.

[37] Niculescu-Mizil A, Caruana R. Predicting good probabilities with supervised learning. In: Proc 22nd Int Conf Mach Learn. 2005:625–632.

[38] Hardt M, Price E, Srebro N. Equality of opportunity in supervised learning. Adv Neural Inf Process Syst. 2016;29:3315–3323.

[39] Chouldechova A. Fair prediction with disparate impact: a study of bias in recidivism prediction instruments. Big Data. 2017;5(2):153–163.

[40] Barocas S, Hardt M, Narayanan A. Fairness and Machine Learning: Limitations and Opportunities. Cambridge, MA: MIT Press; 2023.

[41] Verma S, Rubin J. Fairness definitions explained. In: Proc Int Workshop Softw Fairness (FairWare). 2018:1–7.

[42] Mitchell S, Potash E, Barocas S, D’Amour A, Lum K. Algorithmic fairness: choices, assumptions, and definitions. Annu Rev Stat Appl. 2021;8:141–163.

[43] Platt JC. Probabilistic outputs for support vector machines and comparison to regularized likelihood methods. In: Smola A, Bartlett P, Schölkopf B, Schuurmans D, eds. Advances in Large Margin Classifiers. MIT Press; 1999:61–74.

[44] Zadrozny B, Elkan C. Transforming classifier scores into accurate multiclass probability estimates. In: Proc 8th ACM SIGKDD Int Conf Knowl Discov Data Min. 2002:694–699.

[45] Guo C, Pleiss G, Sun Y, Weinberger KQ. On calibration of modern neural networks. In: Proc 34th Int Conf Mach Learn. 2017;70:1321–1330.

[46] Pleiss G, Raghavan M, Wu F, Kleinberg J, Weinberger KQ. On fairness and calibration. Adv Neural Inf Process Syst. 2017;30:5680–5689.

[47] Vickers AJ, Elkin EB. Decision curve analysis: a novel method for evaluating prediction models. Med Decis Making. 2006;26(6):565–574.

[48] van de Sande D, van Genderen ME, Huiskens J, Gommers D, van Bommel J. Moving from bytes to bedside: a systematic review on the use of artificial intelligence in the intensive care unit. Intensive Care Med. 2021;47(7):750–760.

[49] Marafino BJ, Park M, Davies JM, et al. Validation of prediction models for critical care outcomes using natural language processing of electronic health record data. JAMA Netw Open. 2018;1(8):e185097.

[50] Pfohl SR, Foryciarz A, Shah NH. An empirical characterization of fair machine learning for clinical risk prediction. J Biomed Inform. 2021;113:103621.

[51] Paulus JK, Kent DM. Predictably unequal: understanding and addressing concerns that algorithmic clinical prediction may increase health disparities. NPJ Digit Med. 2020;3(1):99.

[52] Chen IY, Joshi S, Ghassemi M, Ranganath R. Treating health disparities with artificial intelligence. Nat Med. 2020;26(1):16–17.

[53] Barnato AE, Alexander SL, Linde-Zwirble WT, Angus DC. Racial variation in the incidence, care, and outcomes of severe sepsis: analysis of population, patient, and hospital characteristics. Am J Respir Crit Care Med. 2008;177(3):279–284.

[54] Prescott HC, Angus DC. Enhancing recovery from sepsis: a review. JAMA. 2018;319(1):62–75.

[55] Lasser KE, Himmelstein DU, Woolhandler S. Access to care, health status, and health disparities in the United States and Canada: results of a cross-national population-based survey. Am J Public Health. 2006;96(7):1300–1307.

[56] Finlayson SG, Subbaswamy A, Singh K, et al. The clinician and dataset shift in artificial intelligence. N Engl J Med. 2021;385(3):283–286.

[57] Subbaswamy A, Schulam P, Saria S. Preventing failures due to dataset shift: learning predictive models that transport. In: Proc 22nd Int Conf Artif Intell Stat (AISTATS). 2019;89:3118–3127.

[58] Nestor B, McDermott MBA, Boag W, et al. Feature robustness in non-stationary health records: caveats to deployable model performance in common clinical machine learning tasks. In: Proc Mach Learn Health (ML4H) at NeurIPS. 2019;116:381–405.

[59] Char DS, Shah NH, Magnus D. Implementing machine learning in health care — addressing ethical challenges. N Engl J Med. 2018;378(11):981–983.

[60] Ghassemi M, Naumann T, Schulam P, et al. A review of challenges and opportunities in machine learning for health. AMIA Summits Transl Sci Proc. 2020;2020:191–200.

[61] Shickel B, Tighe PJ, Bihorac A, Rashidi P. Deep EHR: a survey of recent advances in deep learning techniques for electronic health record (EHR) analysis. IEEE J Biomed Health Inform. 2018;22(5):1589–1604.

[62] Wexler J, Pushkarna M, Bolukbasi T, Wattenberg M, Viégas F, Wilson J. The what-if tool: interactive probing of machine learning models. IEEE Trans Vis Comput Graph. 2020;26(1):56–65.

[63] Kaur H, Nori H, Jenkins S, Caruana R, Wallach H, Wortman Vaughan J. Interpreting interpretability: understanding data scientists’ use of interpretability tools for machine learning. In: Proc CHI Conf Hum Factors Comput Syst. 2020:1–14.

[64] Goldstein BA, Navar AM, Carter RE. Moving beyond regression techniques in cardiovascular risk prediction: applying machine learning methods. Eur Heart J. 2017;38(23):1805–1814.

[65] Subbaswamy A, Saria S. From development to deployment: dataset shift, causality, and shift-stable models in health AI. Biostatistics. 2021;22(4):1085–1095.

